# Association of MRI-measured cerebral ventricular volume with APOE ε4 genotype, cerebrospinal fluid biomarkers (Aβ42 and Tau) and neuropsychological measures in Alzheimer’s disease: A Systematic Review

**DOI:** 10.1101/2020.12.23.20248759

**Authors:** Albert Dayor Piersson, Mazlyfarina Mohamad, Fadilah Rajab, Subapriya Suppiah

## Abstract

**Rationale and Objectives:** Although neuroimaging studies suggest that the cerebral ventricle is independently associated with APOE ε4, cerebrospinal fluid (CSF) biomarkers, and neuropsychological scores in aging and Alzheimer’s disease (AD), there is no formal synthesis of these findings. We summarized the association of ventricular changes with APOE ε4, CSF biomarkers, and neuropsychological measures.

**Materials and Methods:** The Preferred Reporting Items for Systematic reviews and Meta-Analyses guideline was used. PubMed, Scopus, Ovid, Cochrane, and grey literature were searched, and assessment of eligible articles was conducted using the Newcastle-Ottawa Scale.

**Results:** 24 studies met the inclusion criteria. Progressive ventricular volume is increased in AD patients at an average volume of 4.4 – 4.7 cm^3^/ year compared to average volumes of 2.7 – 2.9 cm^3^/ year and 1.1 – 1.4 cm^3^/year for patients with MCI and healthy controls (HCs) respectively. The ventricular volume is estimated to increase by 1.7 cm^3^/year for progression from MCI to AD. APOE ε4 is an independent risk factor for ventricular enlargement in aging and dementia, with AD patients most affected. The combination of CSF Aβ42 with ventricular volume compared to tau is more robust, for tracking the progression of the AD continuum. Further, the combination of ventricular volume with mini-mental state examination (MMSE) scores is the most robust for differentiating AD and MCI from HCs and tracking the progression of the disease.

**Conclusion:** The combination of ventricular volume with APOE ε4, CSF Aβ42, and MMSE scores independently may be potentially useful biomarkers for differentiating and tracking the progression of AD.

## 1. Introduction

The human cerebral ventricular system, formed by the neuroepithelium serves as an essential interconnected network of cavities filled with cerebrospinal fluid (CSF) and sited within the cerebral parenchyma. Its intrinsic ability to provide support and maintenance of brain function is manifested through a wide array of neurometabolites including vitamins, peptides, growth factors, neurotransmitters and extracellular vesicles (1,2). Critical structures of the ventricular system contribute to the mechanical function, for instance, the ventricular wall is a source of neurons and glial cells of the brain, while the neuroepithelium is responsible for the regulation of shape and expansion of the cavities (3,4). Thus, these components and their functions highlights the characteristics of the ventricular system, epitomizes its presence within the cerebrum as invaluable, and demonstrates that the cerebral tissue alone is inadequate for supporting brain function. However, with aging, the glial cells becomes increasingly vulnerable to impairment which may have adverse effect on their homeostatic functions, reducing their ability to offer neuroprotection, and further leading to age-related pathological changes (5). Additionally, aging may cause glial cells to upregulate routes that adversely affect the function of the neurons and synapses, hence actively contributing to age-related neurodegenerative changes, including dementia, of which the most common is Alzheimer’s disease (AD) (6-8). Neurogenic regions implicated in the development of AD i.e. the hippocampus and dentate gyrus are close to the ventricular system, especially the CSF (9). The direct contact of CSF with the brain parenchyma provides a peculiar advantage for the assessment of neurodegenerative changes i.e. AD, in that brain-related biochemical changes can be assessed via AD-related validated biomarkers i.e. CSF amyloid-β42 (Aβ42) which depicts brain amyloidosis, including tau which although not specific to AD, identifies with neuronal injury. These biomarkers have been reported to be related to apolipoprotein E epsilon 4 (APOE ε4), the main genetic risk factor for the development of AD (10,11), although age is the strongest risk factor (12-15). Further, they are posited to elicit changes even before the onset of symptoms (16,17). The rate of ventricular volume expansion in itself is reported to be linked with increased AD neuropathological hallmarks, Aβ plaques and tau neurofibrillary tangles (18), and it has been reported that large cerebral Aβ plaques cause obstructions leading to reductions in CSF drainage and mobility, and disruption of CSF biomarkers (19,20). The consequence is an increased ventricular volume and progressive periventricular oedema which is accompanied by surface gliosis and cognitive impairment (4,19,20).

The Diagnostic and Statistical Manual of Mental Disorders (DSM–5) (21) acknowledges the contribution of genetic testing for assessing AD genetic risk factor, neuropsychological testing for assessing cognitive domain and the integral role of CSF Aβ and tau, including magnetic resonance imaging (MRI)-detected atrophy in the diagnosis of AD. These biomarkers are also very well recommended in the diagnostic guideline for AD (22,23), given that their combination is posited to improve diagnostic accuracy of early onset of AD and prediction of conversion to the clinical stages of AD (24). However, while systematic reviews have been conducted on CSF Aβ alone (25) or in combination with MRI and positron emission tomography (PET) to evaluate gray matter changes (26), there is less focus on ventricular changes in neurodegeneration, especially in patients with AD. This is further exemplified by systematic reviews that analyzed the relationship between APOE ε4 and gray matter regions, especially the medial temporal lobe structures (27,28). Even though one of these studies summarized the evidence regarding the relationship between ventricular changes and APOE ε4, the evidence presented showed a lack of support for the relationship between the two variables (27), perhaps due to the relatively few number of publications available at the time compared to recent times. Besides, systematic reviews conducted on neuropsychological tests used in the assessment of neurodegenerative changes, especially dementia (29-31) failed to analyze the relationship with cerebral alterations in addition. As research efforts continue to grow to identify the most plausible biomarkers for early diagnosis before clinical symptoms set in, prediction, conversion, and tracking the progression of AD, the current systematic review provides a formal synthesis on the association of ventricular changes with genetic and proteomic fluid biomarkers including neuropsychological measures. Specifically, the following available evidence were summarized: the association of MRI-measured cerebral ventricular volume with (i) APOE ε4 (ii) CSF Aβ42 and Tau, and (iii) neuropsychological measures.

## 2. Methods

### 2.1. Information sources and search strategy

A systematic search of the electronic databases Pubmed (http://www.ncbi.nlm.nih.gov/pubmed/), Scopus (https://www.scopus.com/home.uri), Ovid (http://www.ovid.com/site/index.jsp), Cochrane (https://www.cochranelibrary.com/search) and grey literature (https://www.google.com) was conducted to identify relevant studies till March 19, 2020. Further search was conducted in reference lists of all eligible studies. A combination of words deemed relevant to the study was entered in the electronic search engines. The key words used were: for brain: “intracranial,” OR “brain,” OR “cerebral,” OR “ventricle,” OR “ventricular”; for brain measures: “size,” OR “volume”; for measurement type: “magnetic resonance imaging”, OR “MRI”; for APOE: “APOE,” OR “apolipoprotein,”; for CSF: “cerebrospinal fluid,” OR “CSF,” OR “CSF Aβ42,” OR “CSF amyloid-β,” OR “CSF β-amyloid,” OR “CSF “amyloid beta,” OR “Tau,”; for neuropsychological measures: “neuropsychological measure,” OR “neuropsychiatric measure,” OR “neuropsychological testing,” OR “mini-mental state,” OR “MMSE,” OR “Montreal Cognitive Assessment,” OR “MoCA,” OR “Alzheimer’s Disease Assessment Scale-Cognitive Subscale,” OR “ADAS-cog,” OR “Clinical Dementia Rating” OR “CDR,”; for subjects: “Alzheimer’s disease,” OR “AD,” OR “mild cognitive impairment,” OR “MCI,” OR “dementia” OR “demented” OR “aging,” “ageing,” OR “healthy aging,” OR “healthy cognitive aging,” OR “cognitively normal aging,” OR “cognitive aging.”

### 2.2. Study selection

The articles retrieved had to meet the inclusion criteria set forth: (1) human subjects, but healthy (non-demented) and/ or patients with any type of dementia (P; patient population); (2) structural MRI applied (I; Instrument); (3) ventricular alteration was examined (O; outcome 1); (4) APOE was examined (O; outcome 2); (5) CSF Aβ42 and Tau (O: outcome 3); (6) neuropsychological measures (O: outcome 4); (7) only articles written in English; and (8) full text articles only. Articles were excluded if they did not meet any of the above listed criteria.

Initial screening was conducted by selecting articles based on titles and abstracts appearing to meet the inclusion criteria. Further, the full text of all articles identified at this stage were retrieved obtained. When abstract details were inadequate, full text was obtained for further scrutiny. The next screening phase involved additional scrutiny of full text articles in order to ensure that they met the inclusion criteria. Two independently blinded reviewers (A.D.P and M.M) were involved in these stages. A.D.P. is a PhD candidate conducting a study on neuroimaging of cognitive aging. Consensus were often reached in case there is disagreement regarding any articles. To broaden the scope of the search, reference list of all eligible articles were also hand-searched.

### 2.3. Risk of Bias in Individual Studies

Two independent reviewers (A.D.P. and M.M.) evaluated the methodological quality of all eligible studies. The Newcastle–Ottawa Scale (NOS, http://www.ohri.ca/programs/clinical_epidemiology/oxford.asp) for case–control and cohort studies was used to evaluate the risk of bias. This qualitative assessment scale has been widely used in previous systematic reviews (24,32,33). The NOS is based on 3 subcategories: (selection, comparability, and exposure/outcome) with an overall total of 9 indicative of the highest attained methodological quality. A statement in each subcategory is assigned 1 point. For the “comparability” subcategory, studies were checked if they controlled for age and gender with a point of 1 each allocated. Additional point is given in case any other factor was controlled for i.e. education, ventricular location, e.t.c.). The last statement under subcategory “exposure/outcome” was replaced with a statement to find out if data quality was visually assessed (32). Consensus were reached following comparison of results attained by the two reviewers. On the basis of the research design and methodological quality, each eligible study was assigned a level of evidence (LOE), in accordance with the 2005 classification system of the Dutch Institute for Healthcare Improvement CBO (http://www.cbo.nl/Downloads/632/bijlage_A.pdf). Further, to deduce the strength of conclusion, studies were clustered with comparable studies, thereby accounting not only for the study design but also the risk of bias.

### 2.4. Data Items and Collection

Relevant details extracted from each study are presented in tables 1 to 3. Table 1 includes the following details: (1) authors; (2) magnetic field strength; (3) repetition time (TR)/echo time (TE)/inversion time (TI); (4) flip angle (FA); (5) slice thickness (ST); (6) pulse sequence; and (7) measurement and analysis method.

**Table 1.** MRI Parameters and Measurement Techniques

| Authors | Magnet Field Strength (T) | TR/TE/TI (ms) | FA | ST | Pulse Sequence | Measurement and Analysis Method |
| --- | --- | --- | --- | --- | --- | --- |
| (34) | 3.0 | - | - | - | - | Visual Rating |
| (35) | 1.5 | - | - | 1.6 | 3D T<sub>1</sub>-weighted | Volumetric Analysis (SPM) |
| (36) | 2.0 | 1000/8.6/850 | 20<sup>0</sup> | - | 3D T<sub>1</sub>-weighted MP-RAGE | Volumetric/Manual Tracing |
| (37) | 1.5 | 2400/-/1000 | 8<sup>0</sup> | - | 3D T<sub>1</sub>-weighted | Volumetric /Manual Tracing |
| (38) | - | - | - | - | 3D T<sub>1</sub>-weighted | Volumetric (FreeSurfer) |
| (39) | 1.5 | - | - | - | 3D T<sub>1</sub>-weighted | Volumetric Analysis |
| (40) | 1.5 | 15/7 | 15<sup>0</sup> | - | 3D T<sub>1</sub>-weighted | Volumetric Analysis |
| (41) | 1.0 | 11.4/4.4/400 | - | 1.0 | 3D MP-RAGE | Volumetric Analysis/Manual Tracing |
| (42) | 1.5 | 600/30 | - | 5.0 | T<sub>1</sub>-weighted | Volumetric Analysis/Manual Tracing |
| (43) | 0.22 | 500/30 | - | 10.0 | T<sub>1</sub>-weighted | Manual Tracing |
| (44) | 1.0 | - | - | - | - | Manual Tracing |
| (45) | 3.0 | 3.9/9.5 | 12<sup>0</sup> | 1 | SPGR | Volumetric Analysis (FreeSurfer) |
|  | 3.0 | 9.6/3.9 | 12<sup>0</sup> | 1 |  |  |
| (46) | 1.5 | 2000/20 or 100 | - | 5 | CSE Double-Echo | Semiautomated Segmentation Analysis |
| **(47)** | 1.5 | - | - | - | 3D MP-RAGE | Volumetric Analysis |
| **(48)** | 1.5 | 9/4 | 8<sup>0</sup> | - | 3D MP-RAGE | Volumetric Analysis |
| **(49)** | 1.5 | 2400/-/1000 | 8<sup>0</sup> | - | 3D MP-RAGE | Volumetric Analysis |
| **(50)** | - | - | - | - | - | Volumetric Analysis (FreeSurfer) |
| **(51)** | - | - | - | - | - | Volumetric Analysis |
| **(52)** | 1.5 | 2400/1000 | 8<sup>0</sup> | - | 3D MP-RAGE | Volumetric and Surface Analysis |
| **(53)** | 1.5 | 2400/1000 | 8<sup>0</sup> | - | 3D MP-RAGE | Volumetric and Surface Analysis |
| **(54)** | 3.0 | 9.5/3.9 | 12<sup>0</sup> | 1 | SPGR | Volumetric Analysis |
|  |  | 9.6/3.9 | 12<sup>0</sup> | 1 | SPGR |  |
| **(55)** | 1.5 | 2030/3.93 | - | 1 | 3D MP-RAGE | Volumetric Analysis |
| **(56)** | 1.5 | - | - | - | FLAIR | Volumetric Analysis |
| **(57)** | 1.5 | 600/20 | - | - | T<sub>1</sub>-weighted | Volumetric Analysis (Semiautomated image assessment) |

T: Tesla; TR: repetition time; TE: echo time; TI: inversion time; FA: flip angle; ST: slice thickness.

On the other hand, the evidence tables (Tables 2 and 3) includes the following relevant details: following items: (1) authors; (2) patient group (number of participants, gender and age); (3) healthy controls (HC) (number of participants, gender and age); (4) APOE and structural brain alterations; (5) neuropsychological measures and structural brain alterations; (6) CSF biomarkers and structural brain alterations; (7) neuropsychological tools; and (8) remarks. While tables 2 includes only cross-sectional studies, table 3 includes longitudinal studies.

**Table 2.**
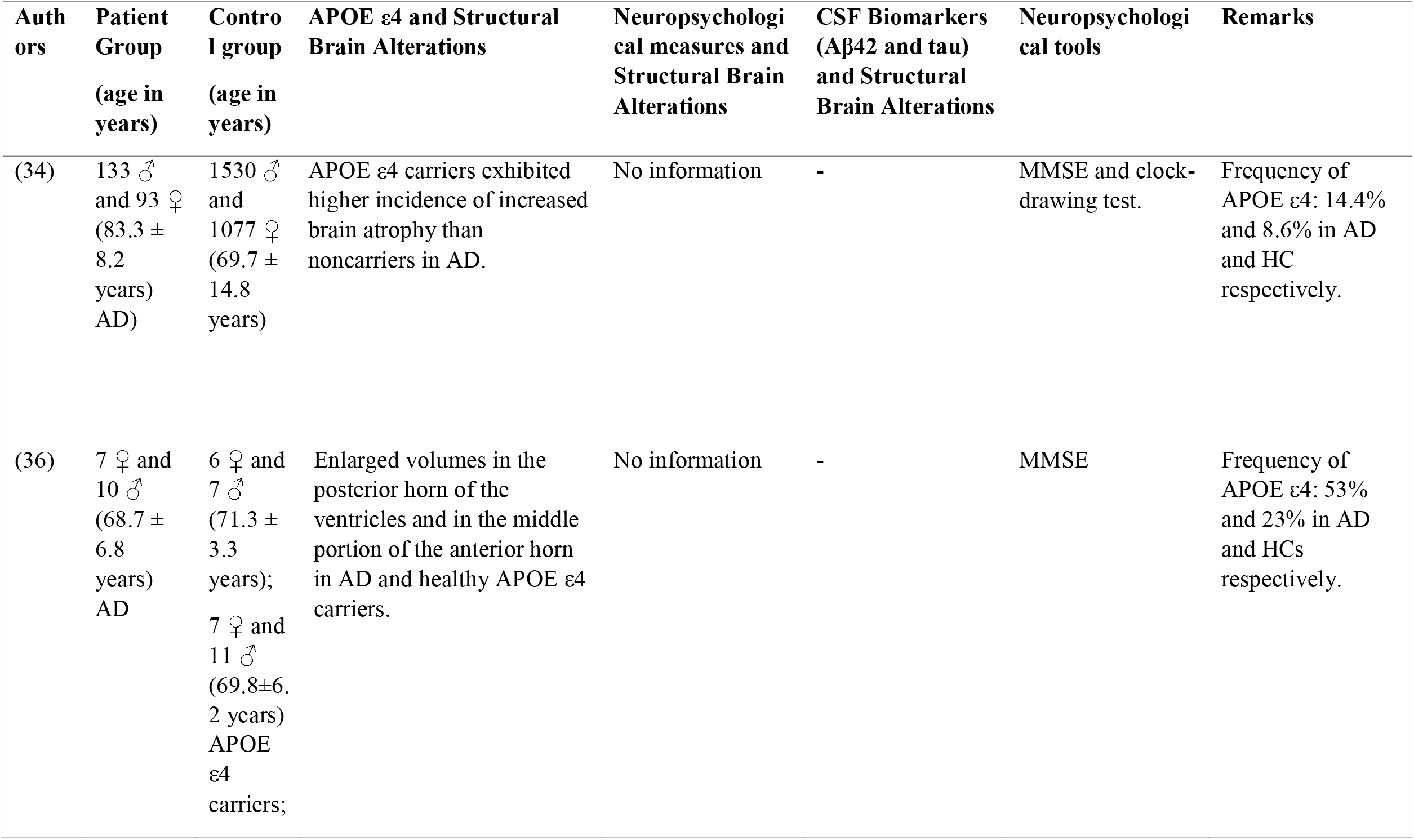
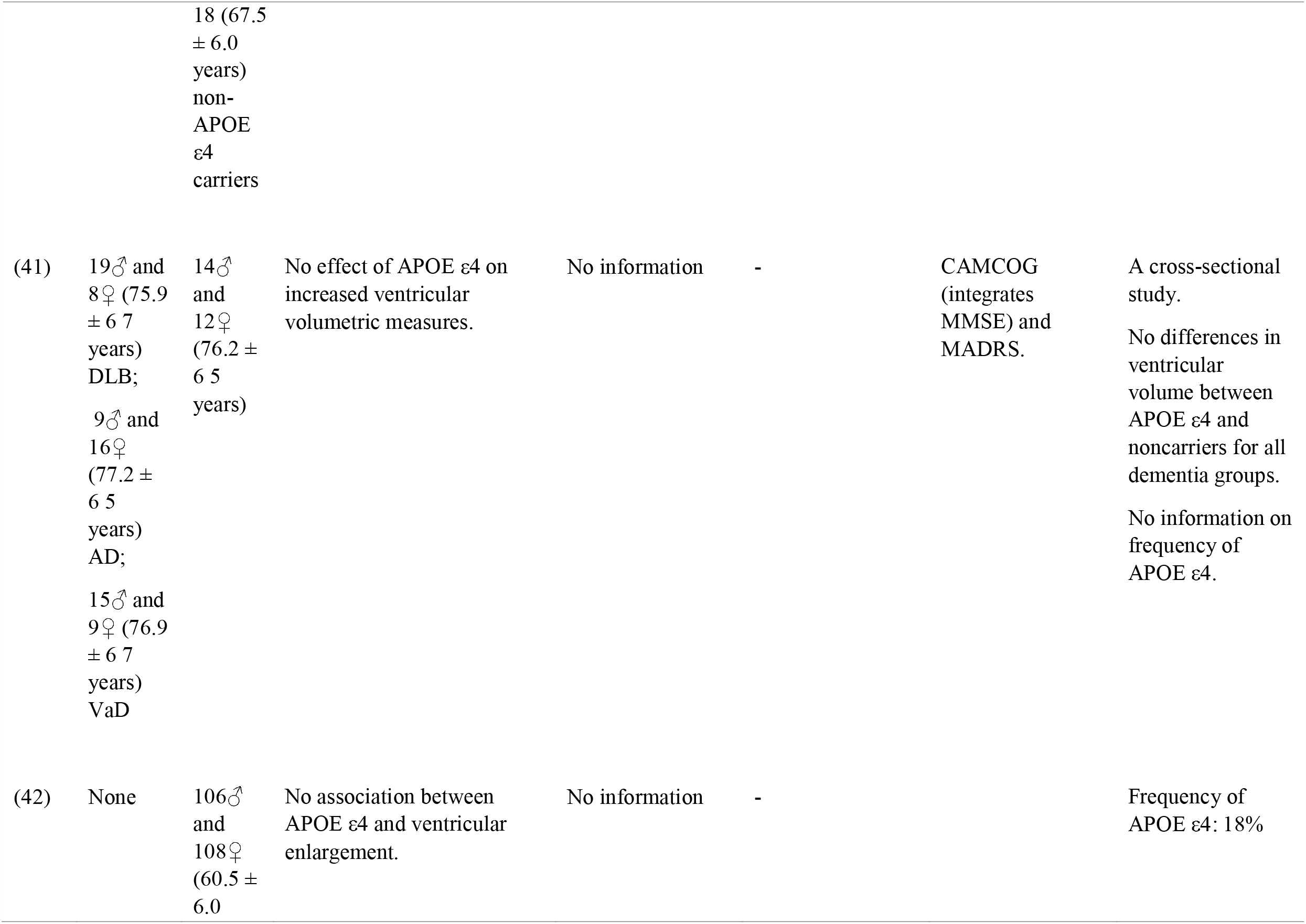
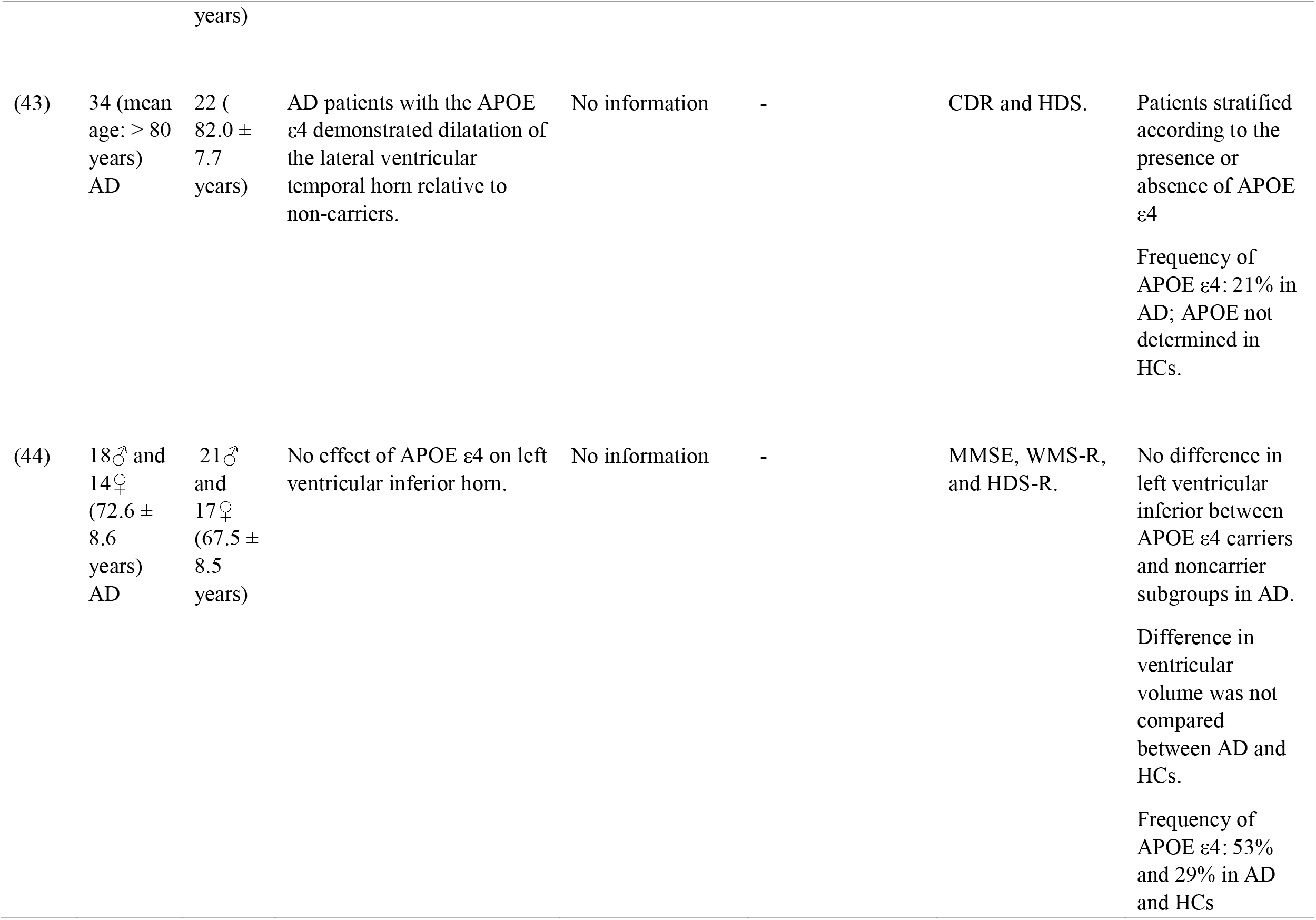
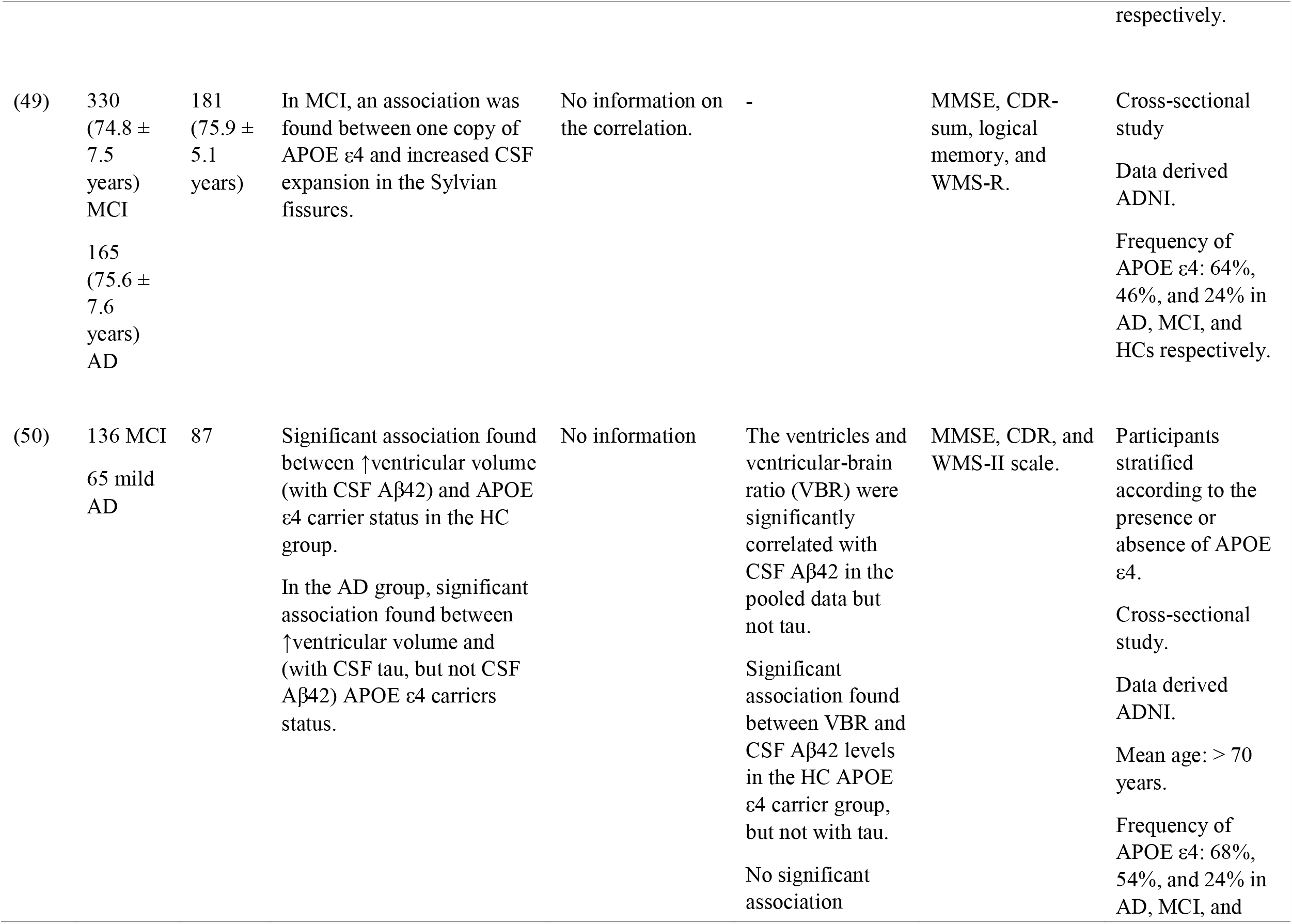
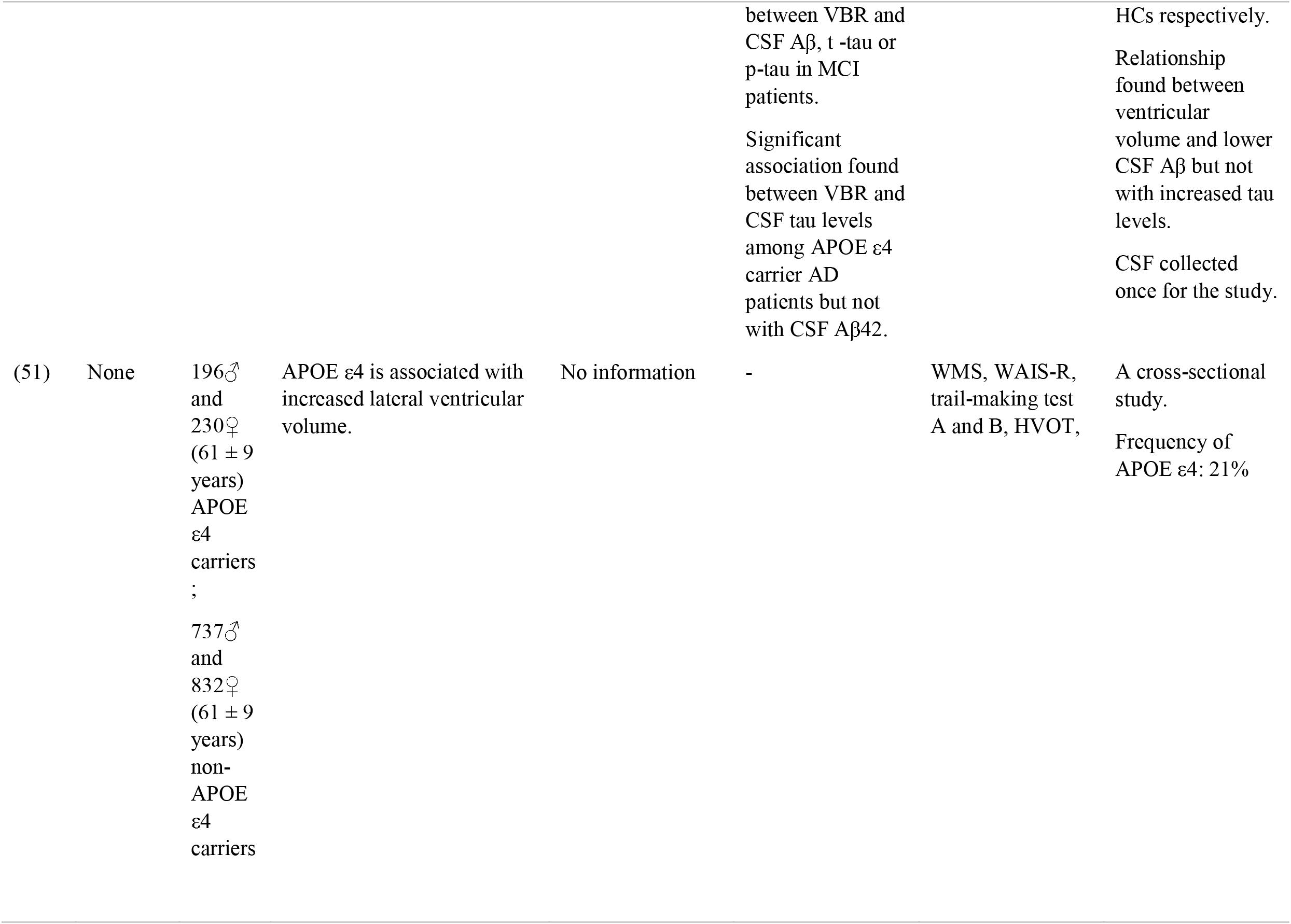
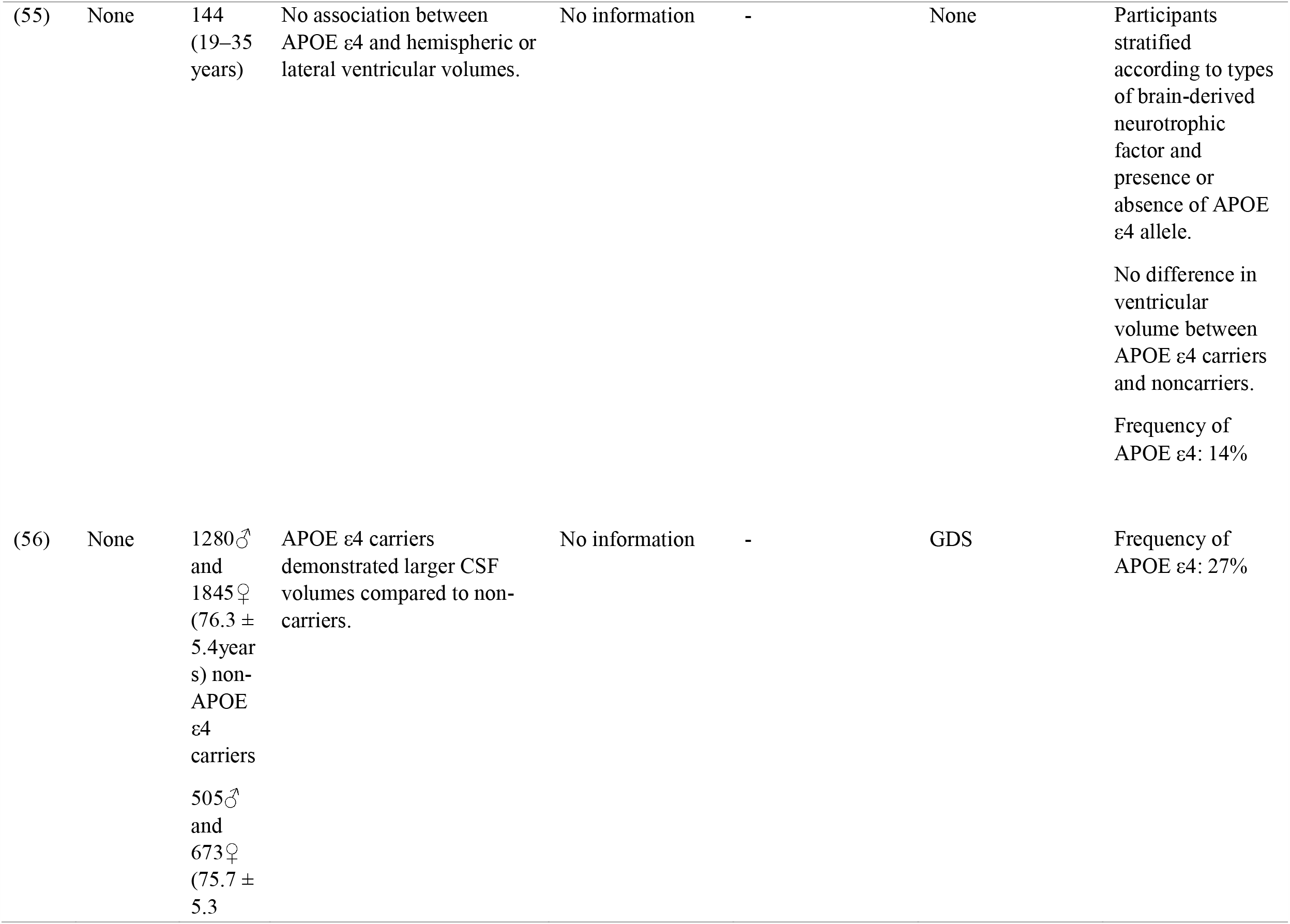
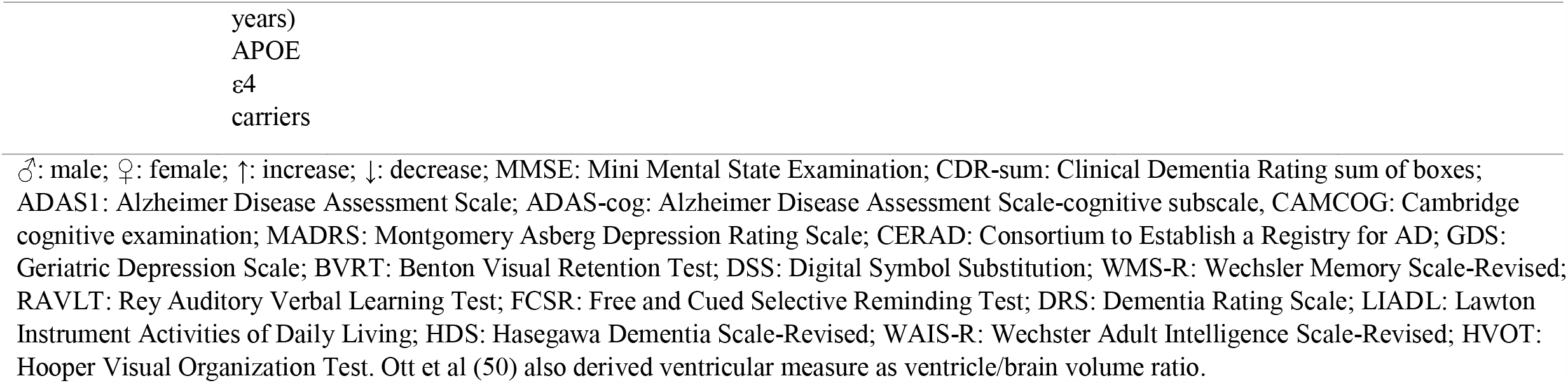
Evidence of the association of MRI-measured cerebral ventricular volume with APOE ε4 genotype, neuropsychological measures, and CSF biomarkers (Aβ42 and Tau) in the cross-sectional studies (n = 11) included in the review.

**Table 3.**
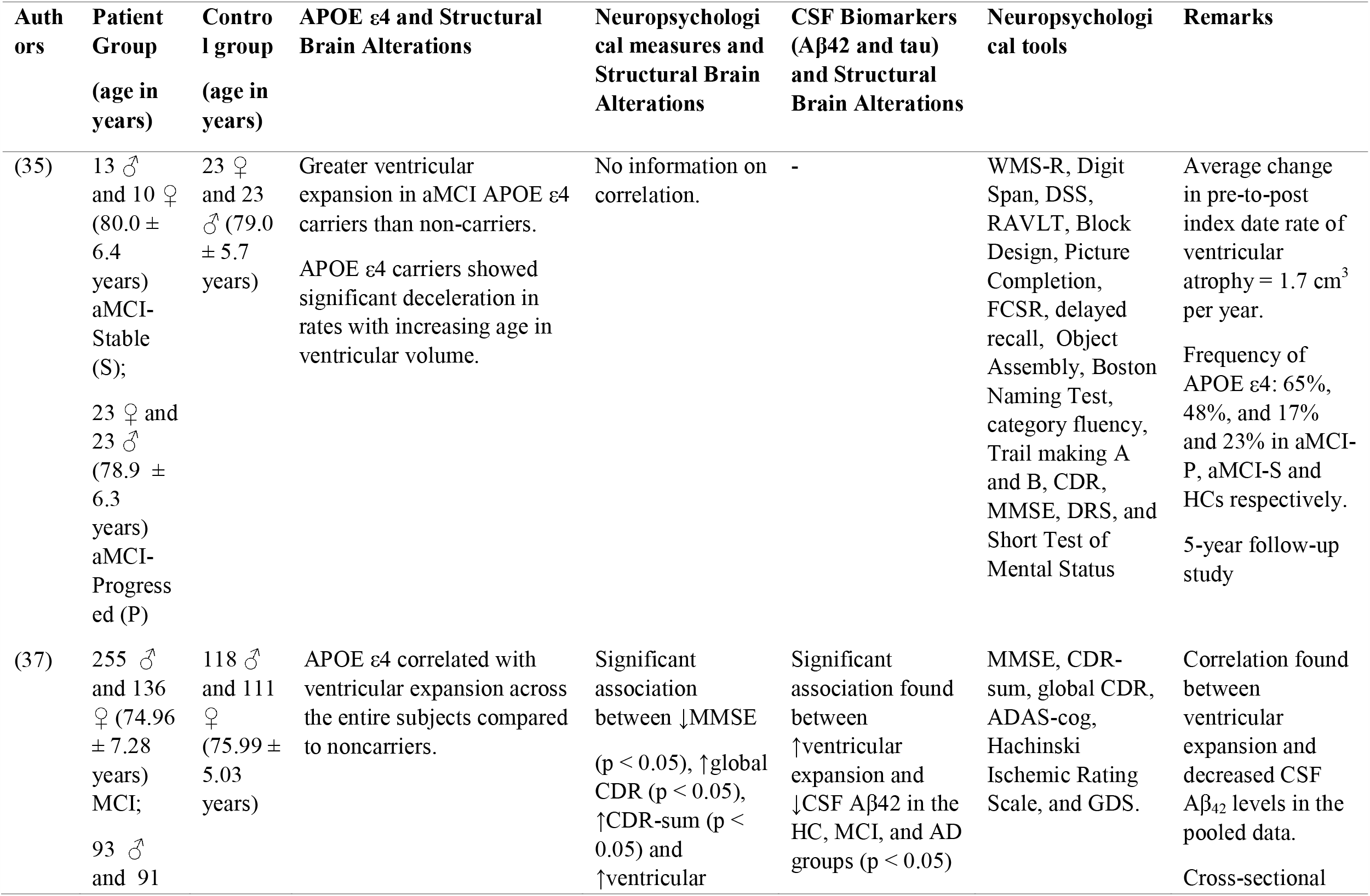
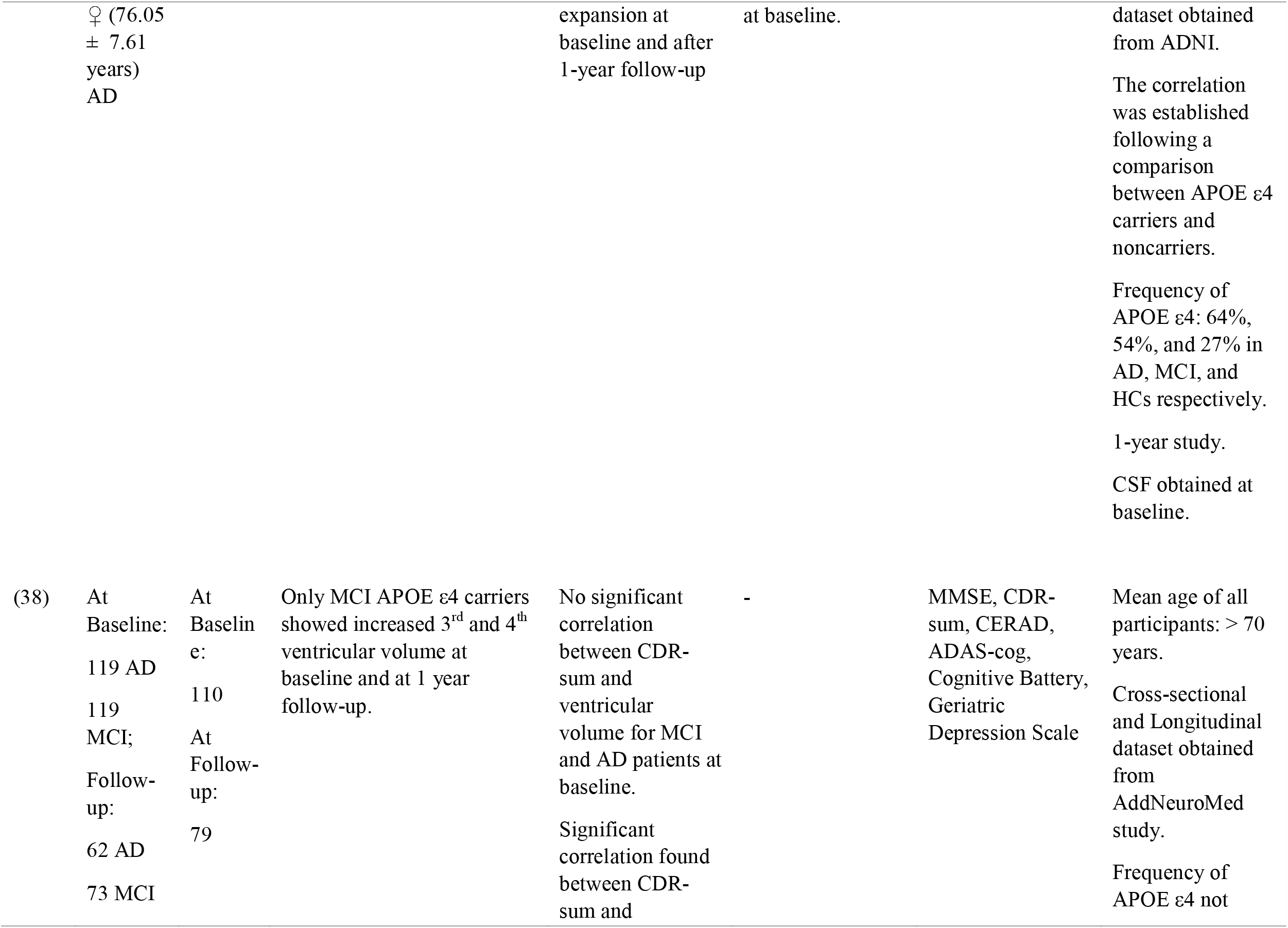
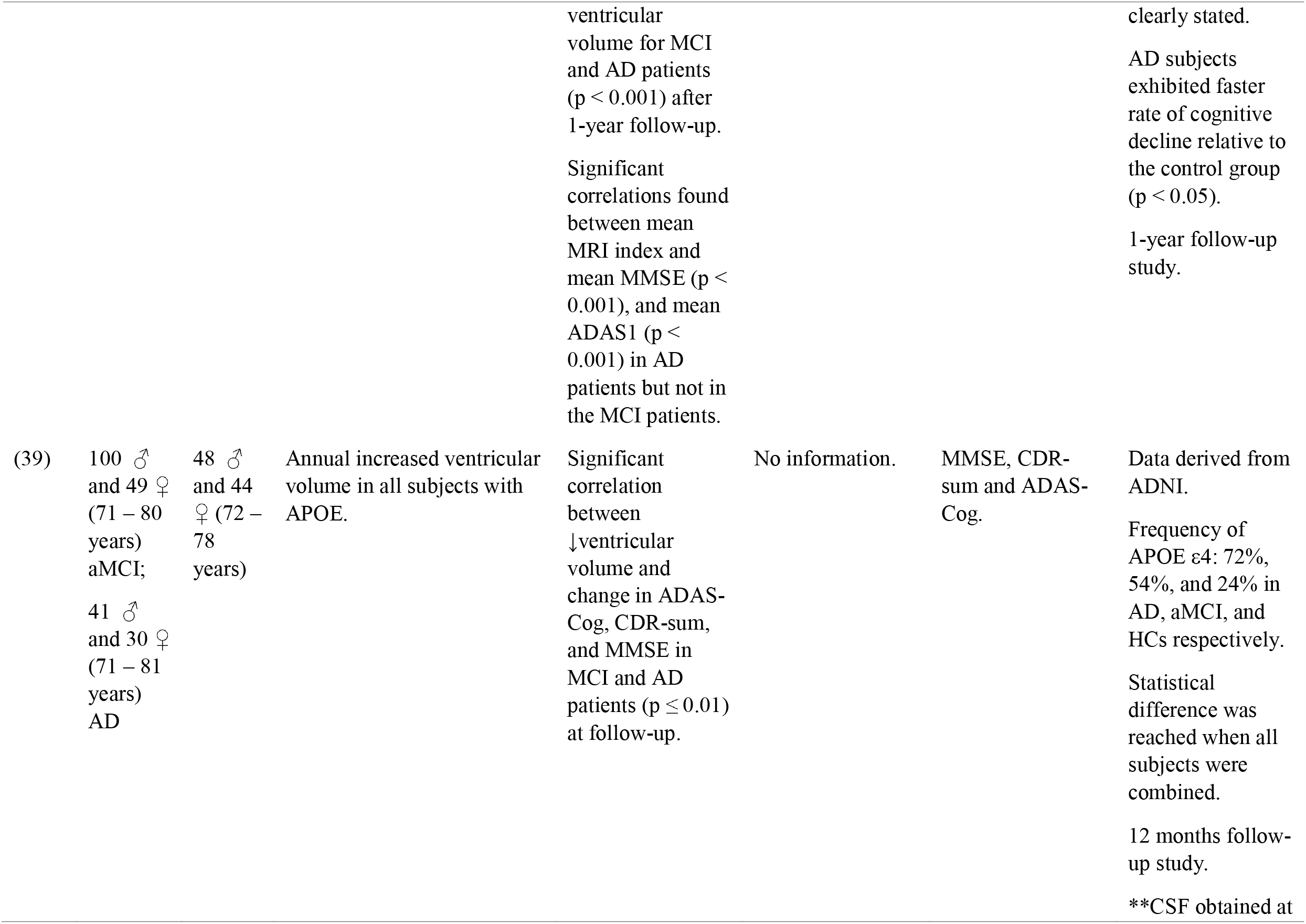
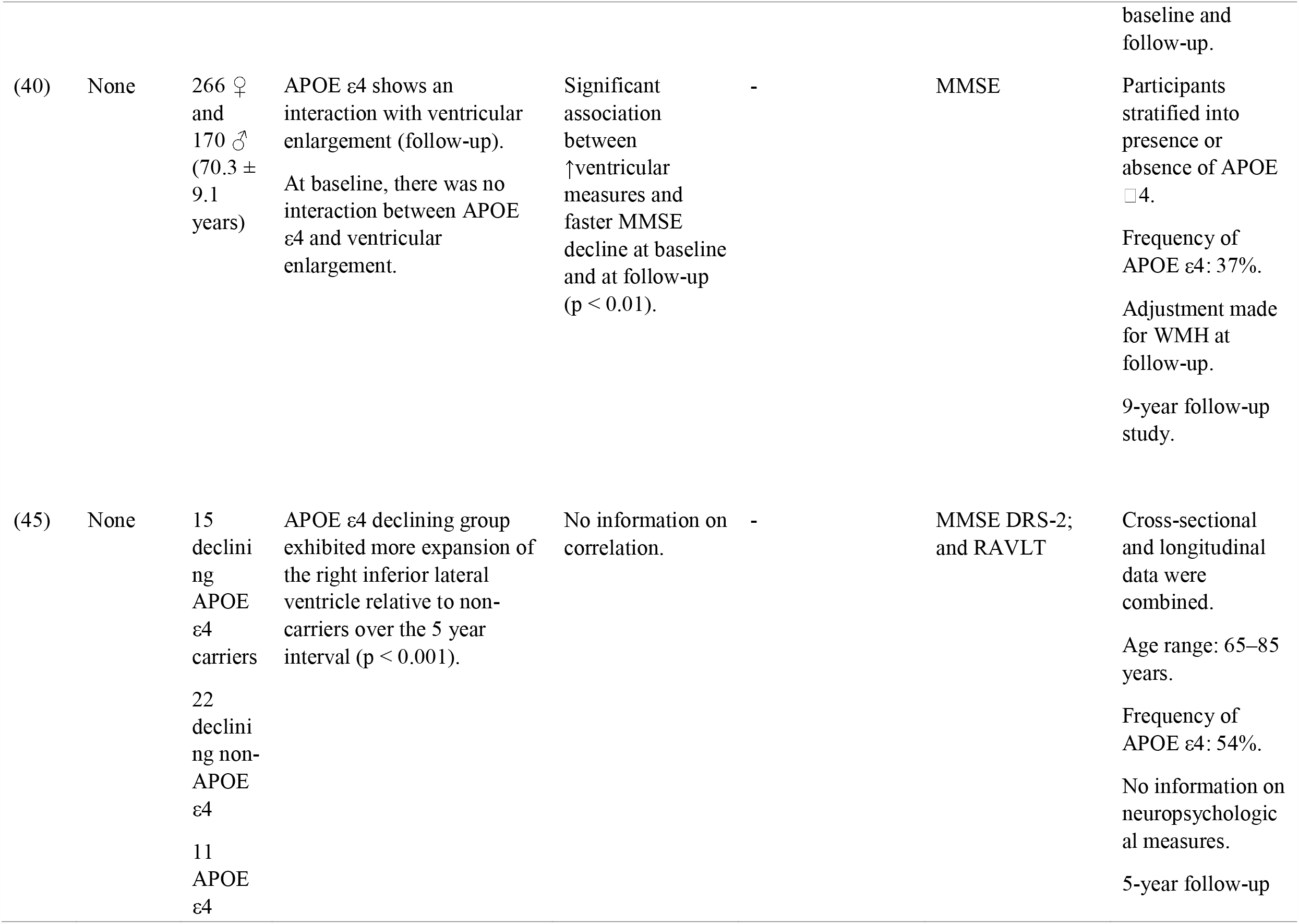
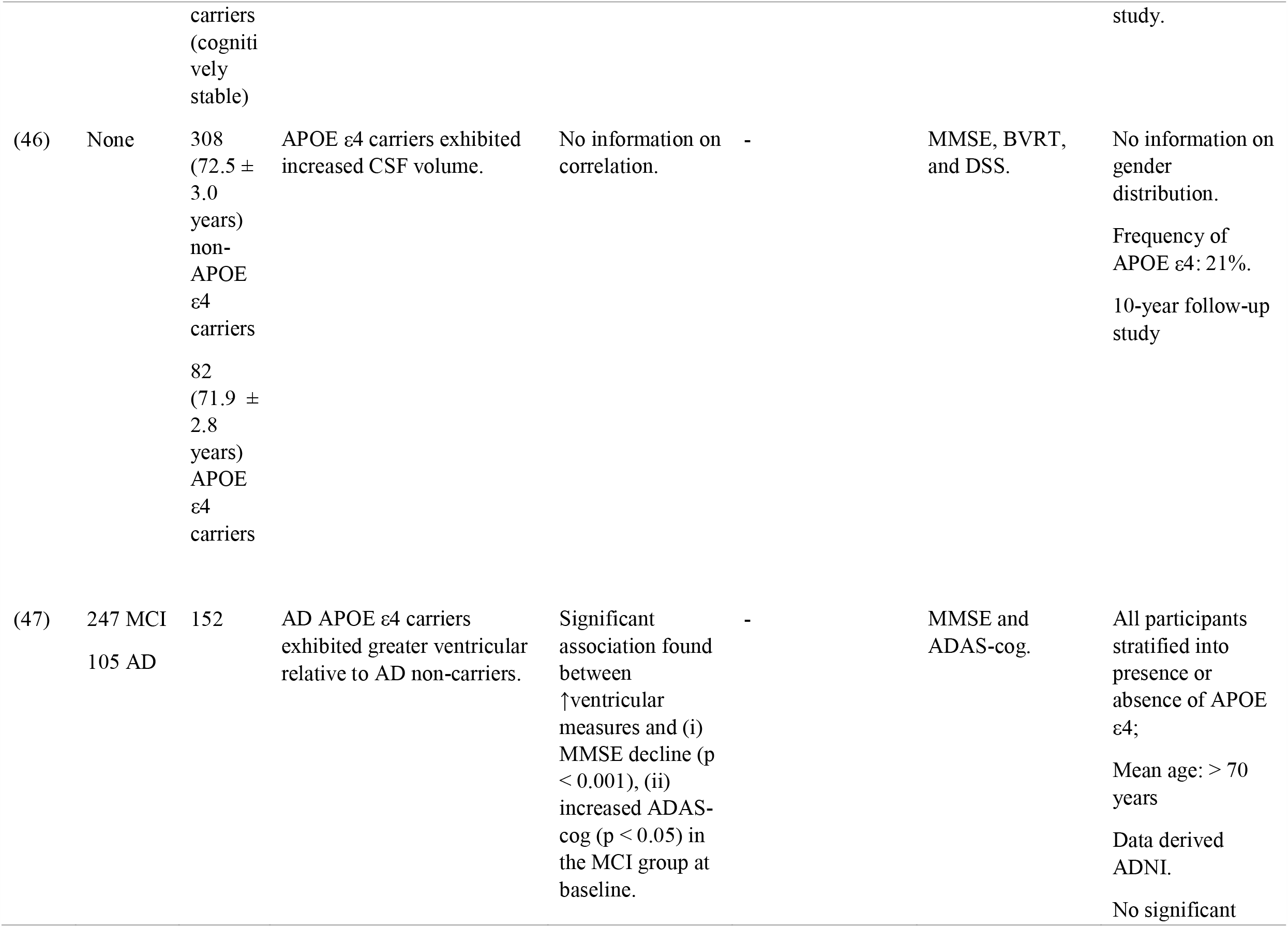
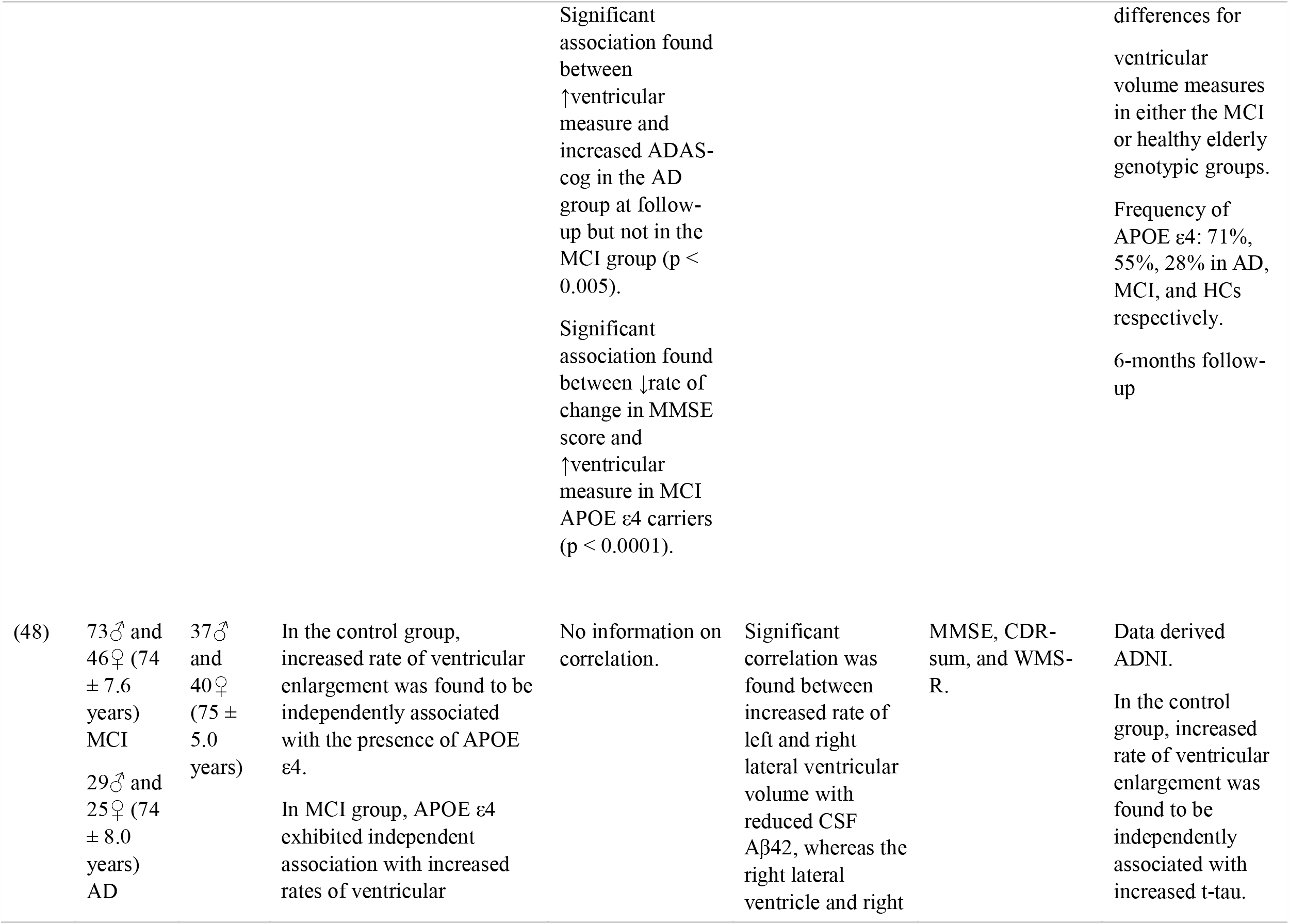
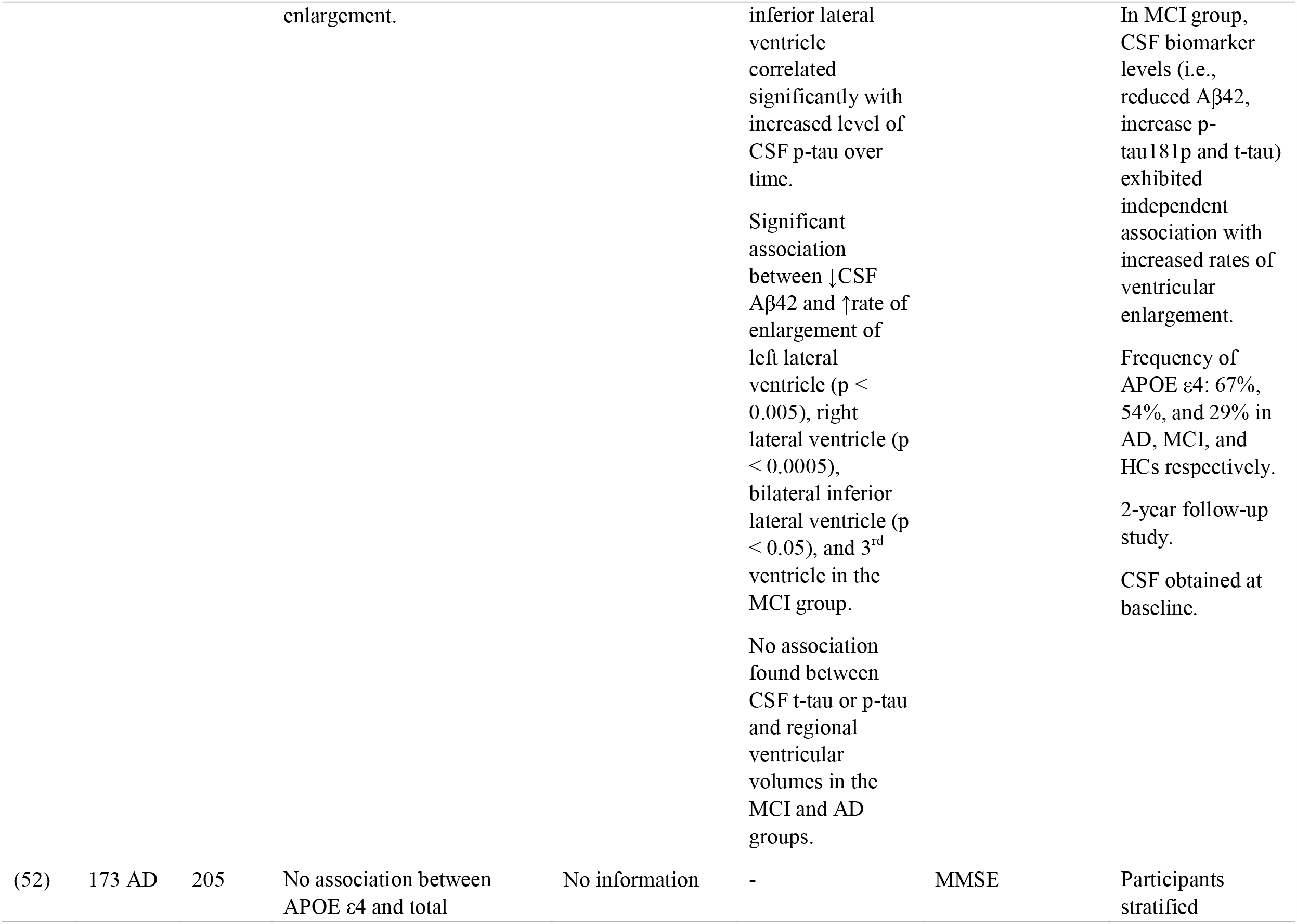
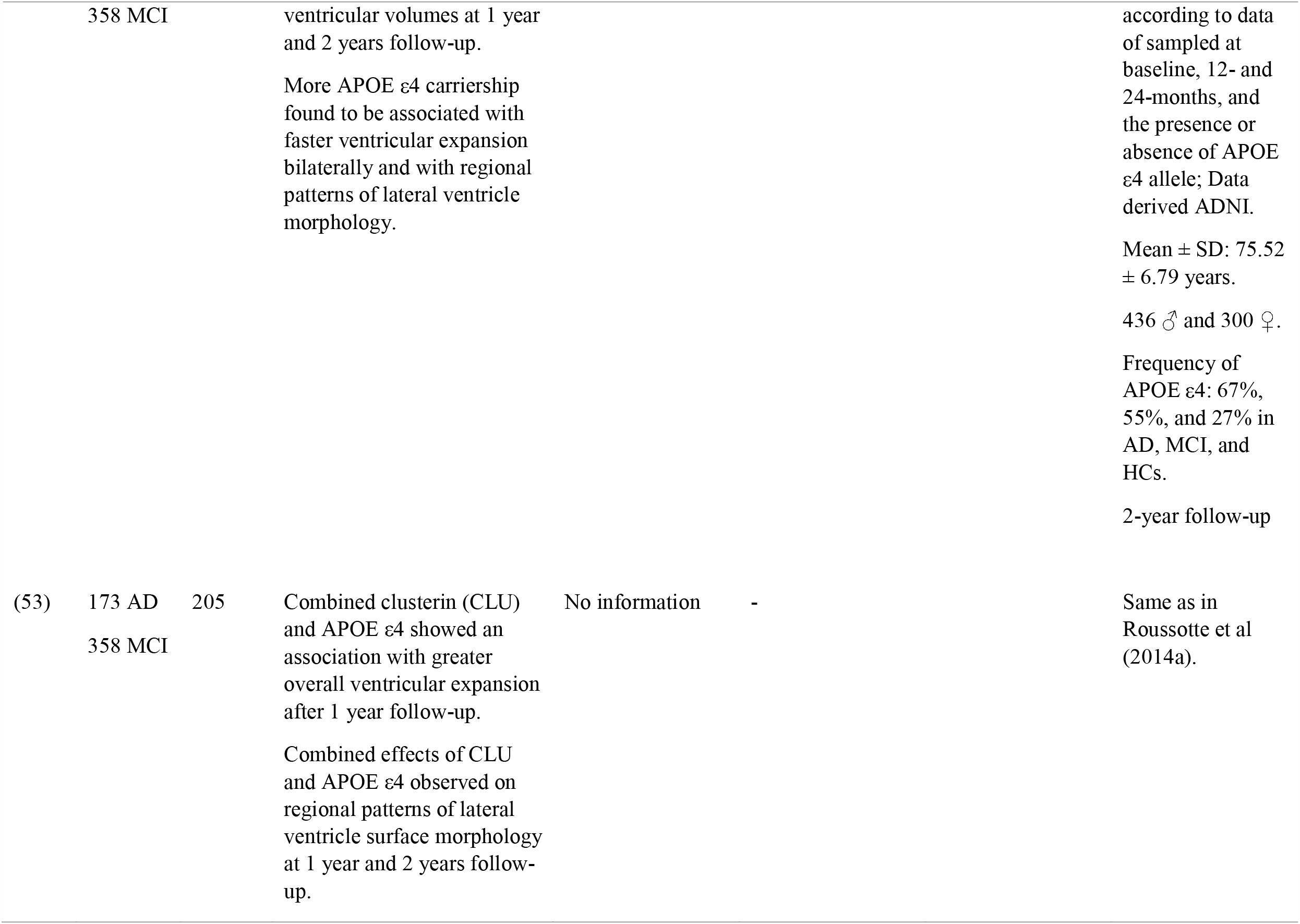
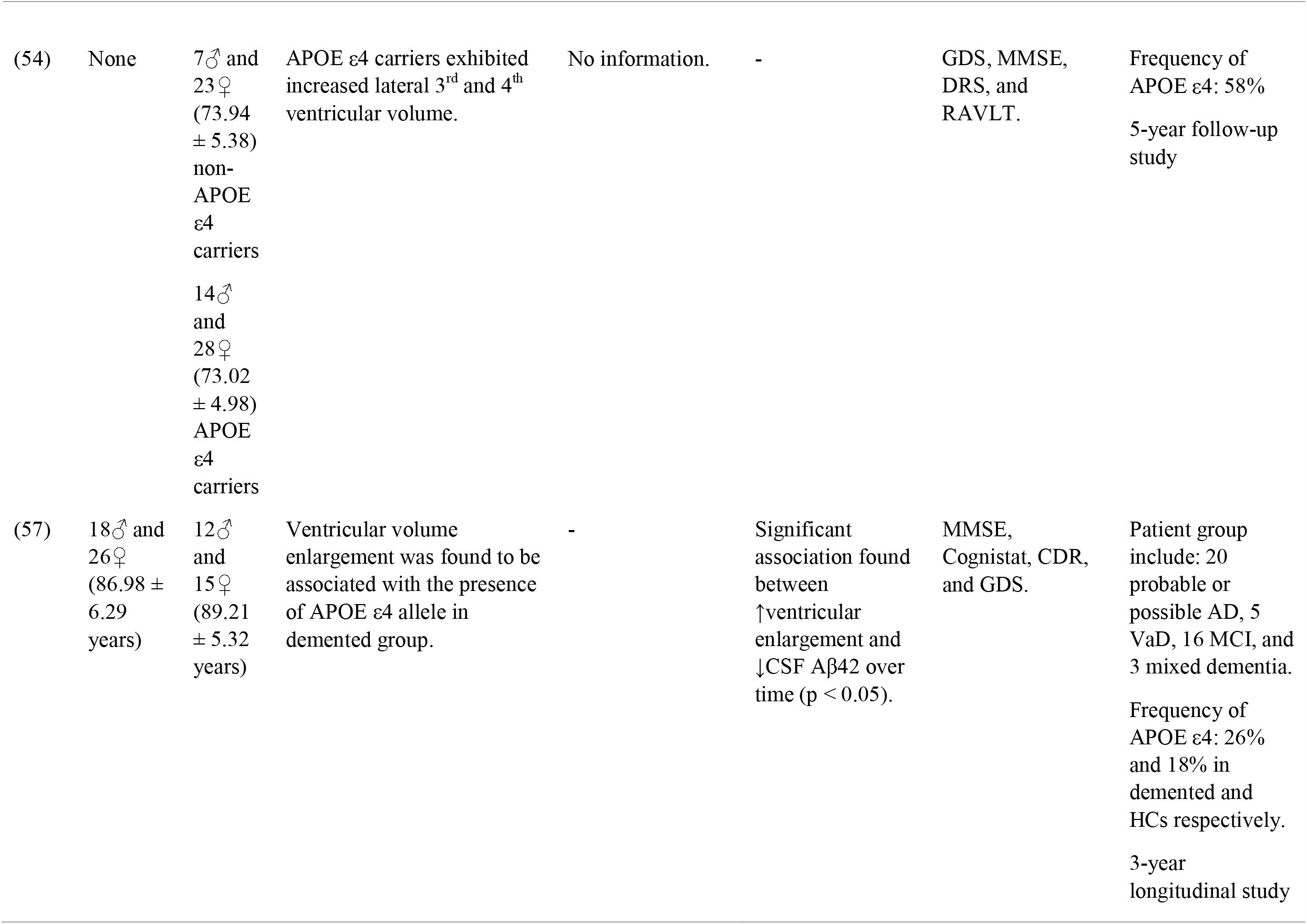
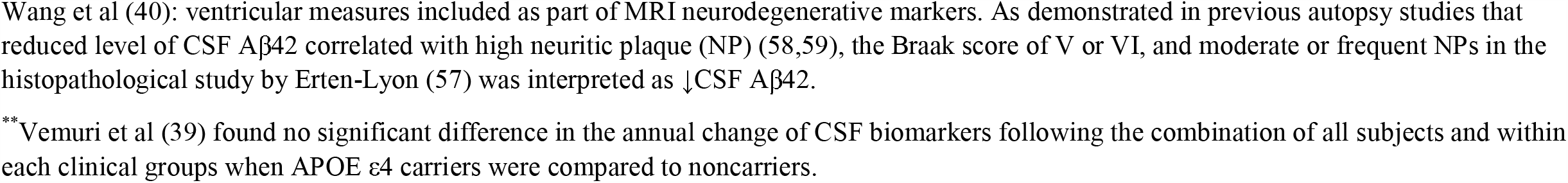
Evidence of the association of MRI-measured cerebral ventricular volume with APOE ε4 genotype, neuropsychological measures, and CSF biomarkers (Aβ42 and Tau) in the longitudinal studies (n = 13) included in the review.

## 3. Results

### 3.1. Study Selection

Figure 1 shows the study selection approach used to identify relevant articles. Initial search yielded a total of 367 articles. Upon removing duplicates, the remaining number of articles amounted to 104. In all, 24 eligible articles met the inclusion criteria. Overall, cerebral ventricular volume was investigated in 21 studies. Three studies (46,49,56) investigated CSF volume. Cerebral ventricular volume was identified as the manual tracing or automated quantitative measure of regional (3^rd^, 4^th^, and lateral) and/or global boundary of the cerebral ventricular size. The CSF volume was identified by either manual or semi/fully automated volumetric measure of the CSF volume.

**Figure 1.**
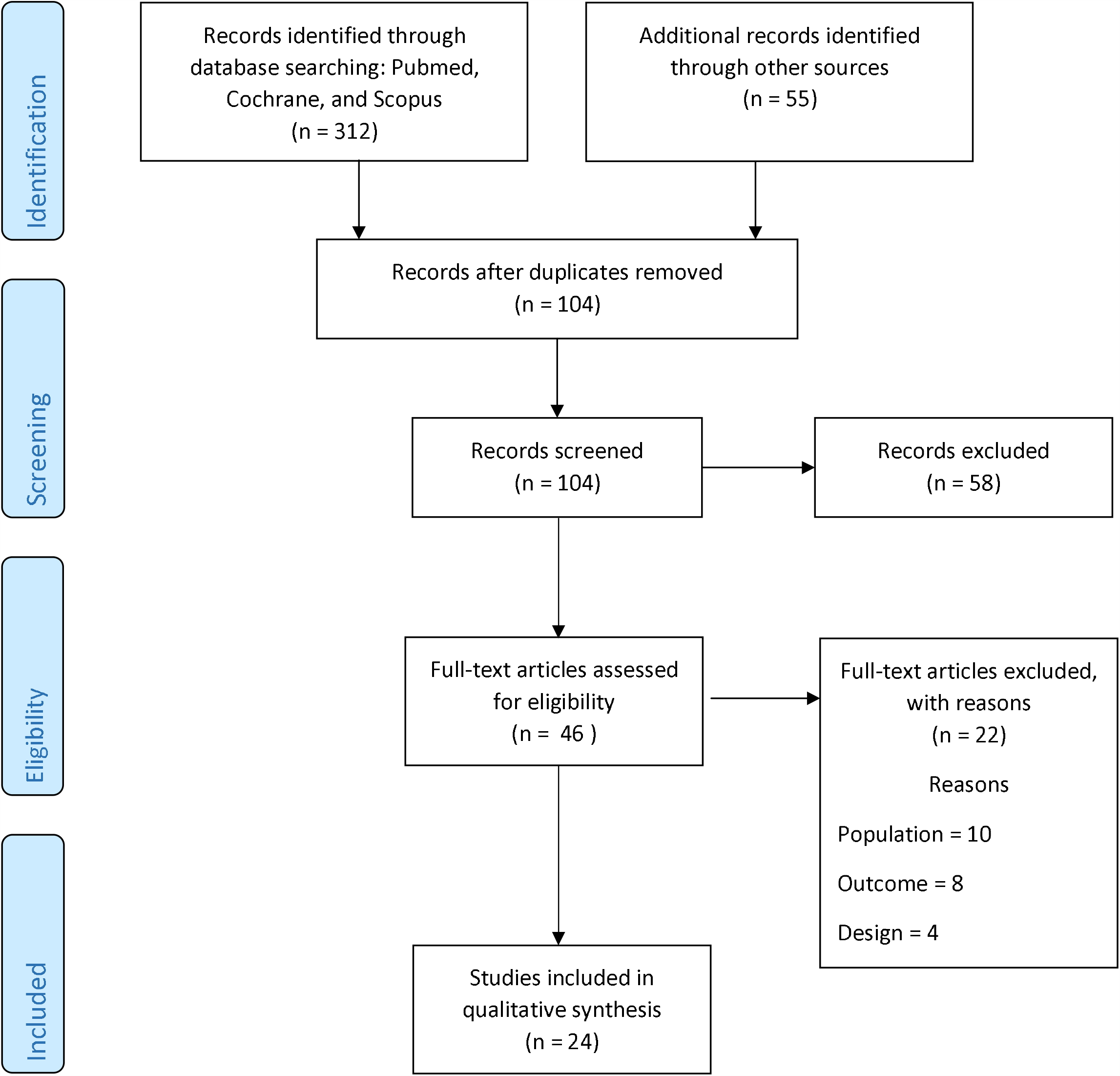
Flowchart for study screening process

### 3.2. Study Characteristics

Of all the eligible studies (n = 24), a total of 14 studies (34-39,43,44,47-49,50,52,53) employed a case-control method, in which patients with AD and/ or MCI were compared with HCs. Further, two studies (41,57) compared a combination of different types of dementia with HCs. Eight studies (40,42,45,46,51,55,54,56) investigated only healthy participants, but compared APOE ε4 carriers to noncarriers. In all, there were 11 cross-sectional studies (Table 2) and 13 longitudinal studies (Table 3). The duration for the longitudinal studies varied from 6 months to 10 years. The mean age of most studies (n = 18) was over 70 years. On the other hand, five studies (34,36,42,44,51) had an average age of participants to be at least 60 years old; whereas only one study had participants between 19 to 35 years (55). Comparatively, the frequency of APOE ε4 in AD surpassed that in patients with MCI, and HC in studies that included all three groups. Two studies did not account for the frequency of APOE ε4 (38,41), and one study did not indicate same in HC participants (43). Only five studies assessed CSF biomarkers (37,39,48,50,57); however, all studies conducted neuropsychological tests.

### 3.3. Risk of Bias Within Studies and Level of Evidence

Table 4 shows the methodological quality for eligible studies, and table 5 shows the LOE. Table 6 shows the strength of conclusion criteria used for clustering of the studies. The LOE scored for all studies was B as they were mainly case-control and cohort studies. Methodological quality assessment for all studies ranged between 5 of 9 to 8 of 9. Most studies lost marks on “representativeness of the cases” and “selection of controls” as it was not clear if the potential for bias existed, and where controls were selected from respectively. In the cohort studies, points were lost for not meeting criteria for “selection of the non-exposed cohort”. Nevertheless, the strength of most studies was that patients were either matched for age and sex with controls or controlled for these variables with one or more additional factors during statistical analysis.

**Table 4:**
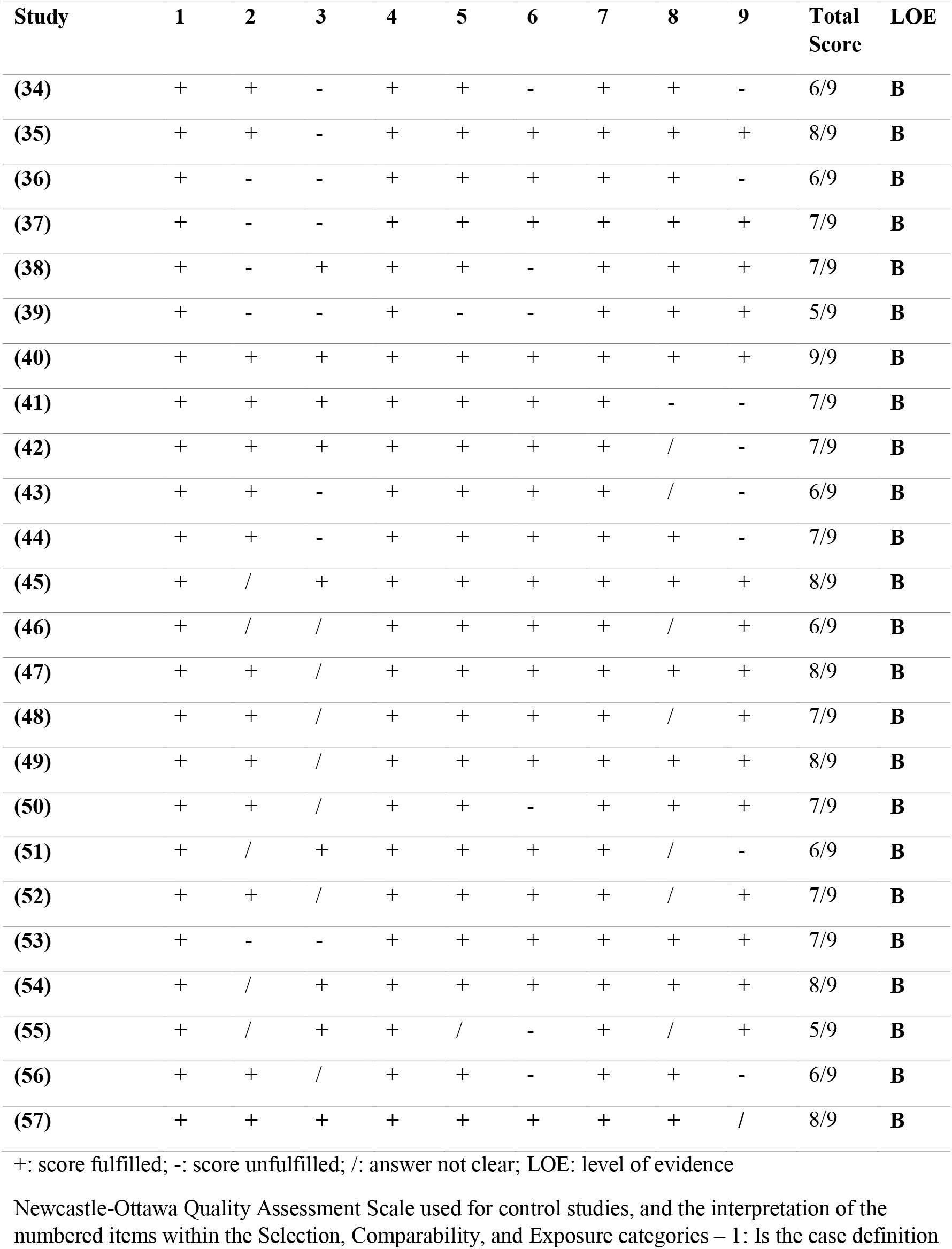
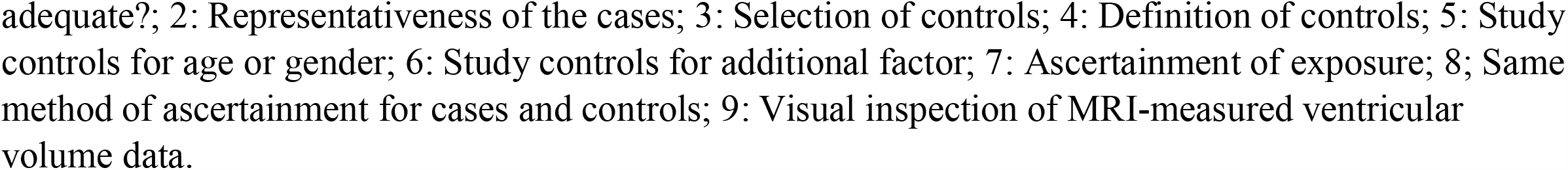
Methodological Quality for Eligible Studies

**Table 5:**
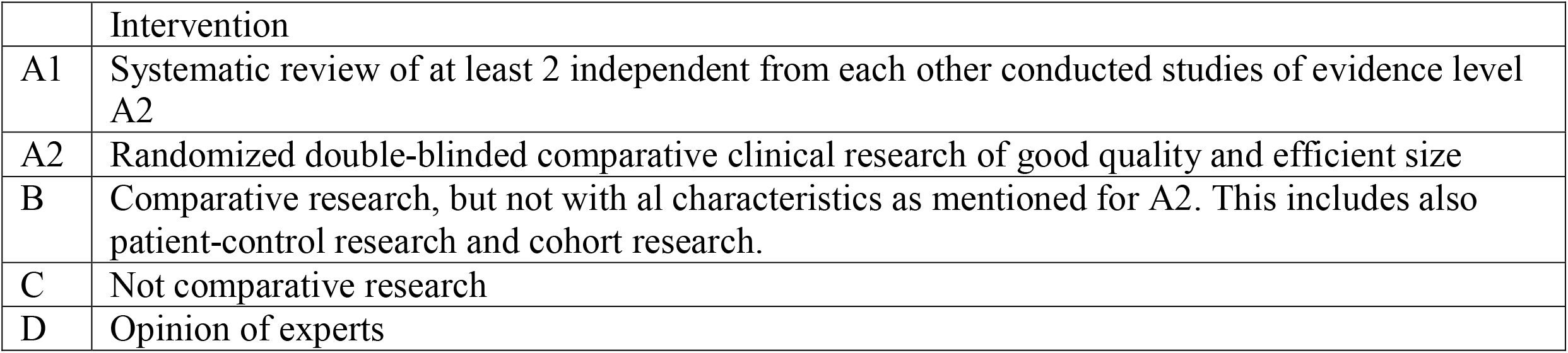
LOE, according to the 2005 classification system of the Dutch Institute for Healthcare Improvement CBO (www.cbo.nl)

**Table 6.** Strength of Conclusion (modified table)

|  | Conclusion based on |
| --- | --- |
| 1 | Research of evidence level A1 or at least 2 independent conducted studies of evidence level A2 |
| 2 | 1 research of evidence level A2 or at least 2 independent conducted studies of evidence level B |
| 3 | 1 research of evidence level B or C |
| 4 | Opinion of experts or Inconclusive or inconsistent results between various studies |

### 3.4. Syntheses of Results

#### 3.4.1. MRI Parameters

As shown in table 1, magnetic field scanner strength ranged between 0.22 – 3.0 T; however most studies used 1.5 T scanners. The most commonly used pulse sequence was 3D MP-RAGE, a T_1-_weighted pulse sequence with a relatively high inherent increased gray and white matter difference in signal-to-noise ratio (SNR) (60). Despite the differences in magnetic field strength, the ventricular CSF T_1_ value does not change with such variation as determined in a previous study that used different magnetic fields (0.2 – 7.0 T) (61). The TR ranged between 3.9 – 2400 ms, and TE ranged between 3.9 – 1000 ms. Since the T_1_ and T_2_ relaxation times of CSF are long, the short TR and TE inherent with MP-RAGE are desirable to reduce the effect of T_2_, thus making CSF to demonstrate hypointense signal on T_1_-weighted images. Other parameter used in MP-RAGE sequence include the inversion recovery time which corresponds to the centre of k-space and leads to increased T_1_-weighted contrast (62). The choice of low FA as used in the studies are also sufficient to shorten the imaging acquisition time which is important for reducing motion artefacts, and for an effective maintenance of the prepared magnetization (63,64). Most studies used a slice thickness that is less than 5 mm, which is commonly used in routine brain imaging. In MRI, slice thickness is one of the parameters that determine spatial resolution and define the voxel slice. Larger voxel size tend to improve the SNR due to increased spins within the volume; however the trade-off is a reduction in spatial resolution and more partial volume effect. Most studies used semi or automatic processing methods. However, manual tracing was performed in 5 studies (36,37,41-43), whereas one study performed visual rating (34). Typically, the “gold standard” for brain volume measurement is manual segmentation (65); however it can be labour intensive when the dataset is large and often requires an expert to perform the trace, thus favouring automated volumetric analysis as a better option.

#### 3.4.2. Cerebral Ventricles and Cerebrospinal Fluid Alterations in APOE ε4 carriers

##### 3.4.2.1. Cerebral Ventricular Volume

Two studies (39,47) accounted for the mean rate of change of the ventricular volume for the HCs, MCI, and AD. Vemuri et al (39) found average rates of 1.1 cm^3^/year, 2.9 cm^3^/year, and 4.4 cm^3^/year for HCs, MCI, and AD respectively. On the other hand, Nestor et al (47) found mean rate of ventricular expansion to be 1.4 cm^3^/year, 2.7 cm^3^/year, and 4.7 cm^3^/year for HCs, MCI, and AD respectively. One study reported that for progression from MCI to AD, ventricular volume expansion was 1.7 cm^3^/year (35).

Nine studies investigated alterations in ventricular volume in APOE ε4 carriers relative to noncarriers and all presented consistent findings of ventricular expansion. Two studies (45,54) investigated only healthy individuals and reported consistent findings of ventricular expansion but in different regions.

The four studies (35,38,39,47) that compared AD and MCI with HCs also presented consistent findings of ventricular expansion in APOE ε4 carriers. For instance, compared to HC group, one study found greater expansion of the inferior lateral ventricle in AD and MCI APOE ε4 carriers at 1 year follow-up relative to baseline (38); another found enlargement in the lateral ventricle in only AD APOE ε4 carriers (47). Furthermore, Jack et al (35) noted greater lateral ventricular expansion in aMCI-progressed APOE ε4 carriers, and Vemuri et al (39) noted that AD APOE ε4 carriers demonstrated the greatest enlargement compared to MCI and HC. The three studies (34,36,43) that compared AD with HCs also reported consistent findings but in different ventricular regions. Compared to the HC group, Chou et al (35) reported enlarged volumes of the posterior horn of the ventricles and in the middle portion of the anterior horn in AD APOE ε4 carriers, whereas Tanaka et al (43) reported dilatation of the lateral ventricular temporal horn in AD APOE ε4 carriers, and Li et al (34) found AD APOE ε4 to demonstrate higher incidence of lateral ventricular expansion.

In conclusion, there is moderate evidence that the average rates of ventricular enlargement in HCs, MCI, and AD is estimated to be 1.1 – 1.4 cm^3^/year, 2.7 – 2.9 cm^3^/year, and 4.4 – 4.7 cm^3^/year for HCs, MCI, and AD respectively (strength of conclusion 2). Further, there is some evidence that for progression from MCI to AD, the mean change in ventricular volume expansion is 1.7 cm^3^/year (strength of conclusion 3). There is moderate evidence of increased ventricular expansion in healthy older adults, MCI, and AD carrying the APOE ε4 allele (strength of conclusion 2). However, there is greater ventricular expansion in patients with AD compared with those with MCI and HC carrying APOE ε4 allele with the lateral ventricle being the most affected (strength of conclusion 2). Of particular importance is that all these studies have an average age of about 70 years, suggesting that APOE ε4 may be conferring more significant effect in older age.

##### 3.4.2.2. Cerebrospinal Fluid Volume

Two studies (46,56) of longitudinal design investigated CSF volume in only healthy individuals and reported consistent findings that APOE ε4 carriers demonstrated increased CSF volume compared to the noncarriers.

In conclusion, there is moderate evidence of increased CSF volume in older healthy adults carrying the APOE ε4 allele (strength of conclusion 2).

#### 3.4.3. Cerebral Ventricles and Cerebrospinal Fluid Alterations Related to APOE ε4 and Dose Effect

##### 3.4.3.1. Cerebral Ventricular Alterations Related to APOE ε4

Ten studies examined the association between ventricular alterations and the presence of APOE ε4 with varied findings in different groups. Chou et al (37) found an association between APOE ε4 and lateral ventricular expansion in all participants. Wang et al (40) performed a longitudinal study in which they investigated only healthy participants and found an association between APOE ε4 and lateral ventricular expansion at follow-up. This is in contradiction with Roussotte et al (52) which found no association between APOE ε4 and total lateral ventricular volumes at follow-up. On the other hand, Roussotte et al (52) found an association between more APOE ε4 carriership and faster ventricular expansion bilaterally and with regional patterns of lateral ventricle morphology. Further, from the study conducted by Wang et al (40), no association was found at baseline, consistent with the findings reported in two cross-sectional studies (42,55) that also enrolled only healthy participants. Note that Sidiropoulous et al (55) included only younger age group (19 – 35 years) and indicated the lateral ventricle. Further, four studies (48,50,51,57) found an association between APOE ε4 and ventricular expansion. The study by Erten-Lyons et al (57) was a clinicopathological correlation study which involved a comparison between patients with different types of dementia (AD, VaD, MCI, and mixed dementia) and HCs.

In the longitudinal studies conducted by Roussotte et al (52) and Roussotte et al (53), relative to whole brain size, total ventricular volume expansion was found to correlate with the number of APOE ε4 alleles carried; and after 1 year follow-up, they found that carrying more APOE ε4 alleles correlated with greater overall ventricular expansion and ventricular surface. Further, at 2-year follow-up, effect sizes were not only larger, but there was more salient relationship demonstrated between the number of APOE ε4 alleles carried and total ventricular expansion (52,53), suggesting a dose effect. However in the study by Roussotte et al (53), no association was found between the number of APOE ε4 alleles and ventricular surface at 2 years follow-up. Note that these two studies emanated from the same research group. Further, Roussotte et al (53) examined the combined effect of APOE ε4 and clusterin (CLU) on ventricular volume expansion and its surface morphology, whereas Roussotte et al (52) exempted CLU. The CLU gene (C allele at the rs11136000 locus) is considered the 3^rd^ strongest genetic risk factor for late onset AD after APOE ε4 allele and the much rarer variant in TREM2 (triggering receptor expressed on myeloid cells 2) (64).

In conclusion, there is inconclusive evidence regarding the association between APOE ε4 and ventricular expansion in the cross-sectional studies found (strength of conclusion 4). On the other hand, moderate evidence show an association between APOE ε4 and ventricular expansion in longitudinal studies (strength of conclusion 2). Consistently all longitudinal studies demonstrated and association between APOE ε4 and greater ventricular expansion on follow-up brain imaging.

##### 3.4.3.2. Cerebral Ventricular Alterations Related to APOE ε4 Dose Effect

Two cross-sectional studies (41,44) reported the absence of an association between APOE ε4 and ventricular volume. Barber et al (41) compared a combined group having DLB, AD, VaD with HCs and found no APOE ε4 effect on increased ventricular volume (lateral and 3^rd^ ventricles). On the other hand, Yoshida et al (44) compared AD with HCs and found no APOE ε4 effect specifically on the left ventricular inferior horn. A key point about the latter study is that no difference in left ventricular inferior measures was found between APOE ε4 carriers and noncarrier subgroups in AD.

In conclusion, there is moderate evidence regarding the absence of APOE ε4 dose effect on ventricular expansion in cross-sectional studies (strength of conclusion 2).

##### 3.4.3.3. Cerebrospinal Fluid Alterations Related to APOE ε4 carriers

Only one study examined the relation between APOE ε4 and CSF (49). They found an association between one copy of APOE ε4 and increased CSF expansion in the Sylvian fissures, a region that provides a clear separation of the frontal lobe and parietal lobe superiorly from the temporal lobe inferiorly.

In conclusion, there is some evidence that APOE ε4 is related to increased CSF expansion in older adults (strength of conclusion 3).

##### 3.4.4. Cerebral Ventricles Related to CSF Biomarkers

Although five studies assessed CSF biomarkers, only four studies (37,48,50,57) provided the results of the association between the cerebral ventricular changes and CSF biomarkers.

Notably, the cross-sectional finding by Ott et al (50) overlapped with a longitudinal study’s baseline finding (37) in which increased ventricular volume correlated significantly with reduced CSF Aβ42 in the pooled data (HC, MCI, and AD) but not with tau. On the other hand, ventricular-brain ratio (VBR), a variant of ventricular volume did not show any significant association with CSF Aβ42, t-tau or p-tau in MCI patients (50). Interestingly, in the AD APOE ε4 carriers, significant association was found between VBR and CSF tau levels but not with CSF Aβ42 (50).

At follow-up, in the HCs, significant correlation was found between increased rate of bilateral lateral ventricular volume with reduced CSF Aβ42, whereas the right lateral ventricle and right inferior lateral ventricle correlated significantly with increased level of CSF p-tau (48). Tosun et al (48) and Erten-Lyon et al (57) found significant association between reduced CSF Aβ42 and increased ventricular volume over time in the patient group.

In conclusion, at baseline, there is moderate evidence that increased ventricular volume is significantly associated with reduced CSF Aβ42 in the entire pooled data (HC, MCI, and AD) but not with tau (strength of conclusion 2). In addition there is inconclusive evidence regarding the association between ventricular volume (VBR) and CSF tau levels in the AD continuum (strength of conclusion 4). Furthermore, there is some evidence that ventricular volume (VBR) is correlated with CSF tau levels but not with CSF Aβ42 (strength of conclusion 3).

At follow-up, there is some evidence that increased rate of the ventricular volume is preferentially associated with lower CSF Aβ42 and increased p-tau levels in the HCs, specifically with the right lateral ventricular volume being sensitive to changes in both CSF biomarkers (strength of conclusion 3). There is moderate evidence of the relationship between increased ventricular volume and reduced CSF Aβ42 in the patient group at follow-up (strength of conclusion 2). Additionally, there is some evidence that increased ventricular volume is not associated with neither CSF t-tau nor p-tau levels at follow-up (strength of conclusion 3).

#### 3.4.5. Cerebral Ventricles Related to Neuropsychological Measures

Five studies provided information on the association between the ventricles and neuropsychological measures.

At baseline, Wang et al (40) found positive correlation between cerebral ventricular volume and lower MMSE score in healthy older adults. Nestor et al (47) and Chou et al (37) found that increased ventricular volume correlated significantly with lower scores on MMSE in the AD continuum. Nestor et al (47) also found correlation between ventricular volume and increased ADAS-cog in the MCI group, while Chou et al (37) also found positive correlation between ventricular volume and global CDR. Aguilar et al (38) did not find any significant correlation between ventricular volume and CDR-sum for both MCI and AD patients. This finding was attributed to the overlap in the CDR-sum (and MMSE scores) between the two patient groups. On the contrary, Chou et al (37) found that ventricular volume is correlated with CDR-sum in the AD patients.

At follow-up, two studies found significant correlation between increased ventricular volume and lower score on MMSE in the AD continuum (AD >> MCI) (37,39). Again, two studies found a correlation between increased ventricular volume and increased ADAS-cog in AD patients (39,47). However, for the MCI patients, while Vemuri et al (39) found a positive correlation between ventricular volume and ADAS-cog, Nestor et al (47) found no correlation. Three studies found positive correlation between increased ventricular volume and CDR-sum in the AD continuum (AD >> MCI) (37-39). Wang et al (40) found an inverse correlation between ventricular volume and MMSE score in the healthy older adults, while Chou et al (37) found a positive correlation between ventricular volume and global CDR.

In conclusion, at baseline, there is moderate evidence to suggest that ventricular volume is associated with lower scores on MMSE in the AD continuum (strength of conclusion 2). In addition, there is some evidence that increased ventricular volume is correlated with (i) increased ADAS-cog and increased global CDR in the clinical stages of MCI and AD and, (ii) MMSE decline in healthy aging (strength of conclusion 3). However, it is difficult to establish whether increased ventricular volume is associated with increased CDR-sum or not in the AD continuum (strength of conclusion 4).

At follow-up, there is moderate evidence that increased ventricular volume is associated with lower MMSE score (AD >> MCI), increased ADAS-cog (mainly AD), and CDR-sum (AD >> MCI), with stronger correlations in the AD than the MCI patients (strength of conclusion 2). However, there is inconclusive evidence regarding the relationship between ventricular volume and ADAS-cog in the MCI patients (strength of conclusion 4). There is some evidence that ventricular volume is inversely correlated with MMSE score (strength of conclusion 3), but positively correlated with global CDR (strength of conclusion 3).

## 4. Discussion

The eligible studies varied in magnetic field strength, MRI parameters, pulse sequences, measurement and analysis techniques, as well as study design. We therefore highlight how these factors may have impacted the outcome of the studies. The use of different types of MRI scanner or magnetic field strength (67) could lead to varied results of commonly used MRI-based semi- or fully-automated brain segmentation techniques (67,68). Alterations in brain morphometry may occur following software upgrades with time (69), including varied sample sizes employed in the studies may have also led to larger variations compared to the original effect size of the structural brain alterations studied (70). Importantly, the stratification of the studies into cross-sectional and longitudinal designs allowed for tracking of the progression of the ventricular changes over time using the APOE ε4 allele status, CSF biomarkers, and neuropsychological measures. Several important findings identified are discussed and important conclusions are presented below.

First, we highlight that even at a minimum follow-up period of 6 months, there was evidence of significant progressive changes of the cerebral ventricles and cognitive function along the AD continuum. The mean rates of change in the ventricular volume for the HCs, MCI, and AD were found to be 1.1 – 1.4 cm^3^/year, 2.7 – 2.9 cm^3^/year, and 4.4 – 4.7 cm^3^/year respectively (39,47) demonstrating that patients with AD have the highest propensity for increased ventricular increase. There is also evidence that for progression from MCI to AD, the ventricular volume increased by 1.7 cm^3^/year (35); however further studies may be required to confirm this outcome. The presence of APOE ε4 was found to be profound in individuals with dementia, and most patients with AD demonstrated a relatively higher prevalence of the APOE ε4 allele compared with MCI and HCs in control studies. This supports the long-held belief that APOE ε4 is a risk factor for developing AD. Across studies, there is moderate evidence of increased ventricular expansion not only in healthy older adults, but also demented individuals carrying the APOE ε4 allele. Most importantly, greater ventricular expansion was found to be more prevalent in those with dementia. Comparative studies showed that AD APOE ε4 exhibited the greatest ventricular expansion than MCI and HC APOE ε4 carriers, and the lateral ventricle was identified to be the most affected region. This suggests that the lateral ventricle may be a potentially useful early biomarker to look out for in the AD continuum. In fact, previous studies (71,72) had also proposed the use of volumetric measures of the lateral ventricles as AD biomarkers over certain period of time. A recent study (73) reported that the lateral ventricle is the next brain anatomical structure after the hippocampus is characterized by early deviation of AD from normal aging trajectory prior to the age of 40 years. Therefore it can be speculated that due to the close proximity of the hippocampus to the horns of the lateral ventricle anatomically, the latter may be a highly susceptible region to AD-related neurodegeneration. It is interesting to find that poorer psychomotor speed and working memory performance on Trails B, and poorer recall on visual delayed memory have been reported to be associated not only with reduced cerebral hemisphere volumes, but also with larger peripheral CSF volumes, including the 3^rd^ and lateral ventricular volumes (74).

However, when we dichotomized the studies into cross-sectional and longitudinal designs, it was found that there is inconclusive evidence with respect to the association between APOE ε4 and ventricular expansion in the cross-sectional studies. This is further supported by the lack of APOE ε4 dose effect on ventricular expansion in such studies, suggesting that the early stages of neurodegeneration may be less influenced by APOE ε4-zygosity. Contrarily, in the longitudinal studies, strong evidence shows that there is an association between APOE ε4 and increased ventricular expansion, with concomitant increased APOE ε4 dose effect. This suggests that the presence of APOE ε4 homozygosity confers stronger effect on ventricular changes than in those with heterozygous allele who in turn show more effect than the noncarriers. Indeed a recent longitudinal shows that APOE ε4 homozygous individuals are at an increased risk of faster cognitive decline, especially in episodic memory (75). Longitudinal studies may provide better insight regarding pathophysiological changes underlying the progression of brain atrophy compared to cross-sectional studies. Further, we found moderate evidence supporting the presence of increased CSF volume in older APOE ε4 carriers. This outcome was not found in patients with dementia, likely because most studies rarely account for CSF volumetric measures. We therefore speculate that while increased CSF volume may signify the presence of APOE ε4 in older adults after controlling for age, it may not be a potentially useful AD biomarker. This is buttressed by a recent evidence (76) which found non-significant inverse correlations between ventricular CSF volume and CSF biomarkers. One critical point to note is that in the exception of one study which employed younger adults (55), all the studies had a mean age of at least 60 years old. This suggests that the presence of APOE ε4 may weigh significant deleterious effect on the cerebral ventricular system in the older age group and those with dementia compared to the younger adults. However, more studies involving younger to middle age groups are required to compare how the influence of age across lifespan affect ventricular expansion in APOE ε4 carriers. It is important to point out that several factors can as well interfere with the observed relationship between APOE ε4 and ventricular expansion, and this can include the presence of cerebral microbleeds (77), periventricular hyperintensities (78), increased visceral adipose tissue (79), and osteoporosis (80). Accordingly, these factors to some extent may also exert some effect on the cerebral ventricular system.

It is important to note that as the collection of CSF is usually not feasible for longitudinal population studies, all the studies that analyzed CSF biomarkers obtained CSF at baseline, in the exception of Vemuri et al (39) who obtained additional CSF at follow-up. But yet still, they found no significant difference in the annual change of the levels of CSF biomarkers (39), hence for the longitudinal studies, the analysis were based on baseline CSF biomarkers and ventricular volumes obtained at follow-up. At baseline, we found that the results of the relationship between cerebral ventricular volume and CSF biomarkers shows a stronger preference for Aβ42 compared to tau. At baseline, the pooled data which consisted of the HCs, MCI and AD showed that increased ventricular volume correlated significantly with reduced CSF Aβ42 (but not with tau) in two studies (37,50). On the other hand, ventricular volume correlated positively with CSF tau in AD APOE ε4 carriers (50). This might imply that ventricular volume at the onset may not be sensitive enough, perhaps not increased to the extent that it can interact differentially with CSF Aβ42 for differentiating MCI and AD from HCs, suggesting the need to find out what the outcome may be when gray matter volumes are combined with CSF biomarkers at baseline. However, this correlation as found also supports several evidence in the literature that CSF Aβ42 is a more robust biomarker relative to tau, given its ability to correlate with ventricular volume across all groups. This is evident in the diagnostic guidelines for AD in which reduced levels of CSF Aβ42 is considered an essential AD biomarker indicating brain Aβ amyloidosis (22,23). The lack of relationship between ventricular volume and CSF tau (37,50) and positive relationship between the two variables (50) suggests that their combination may not be specific to AD, in particular. CSF tau in itself is considered to be unspecific to AD, but instead a biomarker of neuronal injury biomarker (22,23), hence it is not surprising to find that its combination with ventricular volume still did not improve its diagnostic performance. Again, the robust performance of CSF Aβ42 was evident at follow-up, where two studies found it to be inversely correlated with the ventricular volume in the patient groups (48,57), suggesting that the combination of these markers may suffice for predicting and tracking the progression of different types of dementia, but more favourably for patients within the clinical stages of MCI and AD. The robustness of CSF Aβ42 have been shown in several studies with evidence that it is able to detect the accumulation of cortical Aβ earlier than Aβ-PET (81,82). However, recent systematic review shows that the predictive accuracy of low levels of CSF Aβ42 for conversion from MCI to AD is low when used as a stand-alone AD biomarker (83), and that it is limited for monitoring disease progression (82,83). Therefore the combination of low CSF Aβ42 with increased ventricular volume may fill this gap. However, other CSF non-Aβ and non-tau have also been proposed for predicting the progression of AD, including for early diagnosis, and they include neurofilament light polypeptide, neurogranin, and visinin-like protein 1 (84). Comparatively, we found that CSF t-tau or p-tau did not correlate with ventricular volume in these same groups at follow-up (48), again supporting the evidence of its lack of specificity for AD. However, there is suggestion that increased rate of the ventricular volume is preferentially associated with lower CSF Aβ42 and increased p-tau levels in the healthy older adults, with the right lateral ventricular volume being more sensitive to both biomarkers.

Interesting results were found regarding the relationship between cerebral ventricular volume and neuropsychological measures. Two studies found an inverse relationship between ventricular volume and MMSE score in the AD continuum at baseline (37,47), while one study found the same relationship in healthy older adults (40). On the whole, MMSE outperformed all the other neuropsychological tools at baseline, thereby demonstrating its superiority in performance compared to ADAS-cog (47) and global CDR (37). The use of MMSE as a stand-alone neuropsychological tool for identifying patients with MCI who may develop dementia is not supported, as revealed in a systematic review (5). Therefore, the combination of ventricular volume with MMSE scores improved its performance in differentiating AD and MCI from HCs at baseline. This approach has also been explored in other cross-sectional studies which found MMSE to be significantly correlated with atrophy of AD-vulnerable regions on MRI (87) and ^18^F-fluorodeoxyglucose-PET in MCI and AD (88). However, it is difficult to establish a statement as to whether increased ventricular volume is associated with increased CDR-sum (37) or not (38) in the AD continuum at baseline. At follow-up, results of the correlations between increased ventricular volume and the three main neuropsychological tools were mainly found in the AD continuum, with stronger relationships found in patients with AD than the MCI patients. Again, inverse relationship was found between ventricular volume and MMSE (37,39). Interestingly, the association of two neuropsychological tools with ventricular volume that were not well-supported at baseline proved supportive at follow-up. This is exemplified by the positive correlation demonstrated by ventricular volume with ADAS-cog (39,47), and CDR-sum (37-39). A previous longitudinal study has also shown the invaluable role of ADAS-cog, suggesting that “orientation” is the only item that is capable of demonstrating detectable cognitive decline in mild AD after 1-year (89). Similarly, the ability of CDR-sum has also been reported to be capable of tracking the progression rate of patients with late MCI and mild AD, with an estimated report of 0.5 points/year and 1.4 points/year respectively (90). Again, we found a weak support for the correlation between ventricular volume and global CDR. Overall, the combination of MMSE, ADAS-cog, and CDR-sum independently with ventricular volume suggests that AD and MCI patients were not only more cognitively impaired, but also more atrophied over time. These combinations can therefore be potentially useful biomarkers for tracking the progression of AD, however there is a stronger support for using the relationship between MMSE and ventricular volume. This is also supported in a recent study which found that the sensitivity of MMSE (∼92%) (but similar to that of ADAS-cog) supersede that of CDR-sum (∼78%) for the identification of changes associated with the clinical stages of AD longitudinally (91).

### 4.1. Strengths and Limitations

Importantly, the present systematic review did not dwell only on the inclusion of cross-sectional studies, but also included longitudinal studies which was instrumental for unraveling the direction of the associations between the variables, and to ascertain the causality: Are cerebral ventricular alterations related to APOE ε4 allele, CSF biomarkers, and neuropsychological measures? The methodological quality of the eligible studies was moderate to good, the LOE scored B as case–control and cohort studies were included. Further, the screening process and scoring were conducted by 2 researchers who were independently blinded. Most studies also controlled for age and gender, which are two key factors that can also have effect on the cerebral ventricular system. The relatively small sample size employed in a few of the studies and disproportionate distribution of gender may be a limitation. In addition, studies employing other neuroimaging techniques were not included as they were thought to be less explored.

### 4.2. Recommendations for further research

Cerebral ventricular measures are infrequently explored in studies interrogating structural brain changes in APOE ε4 carriers who may be cognitively normal or cognitively impaired compared to gray matter regions of the medial temporal lobe. In future, it is important to analyze ventricular measures in addition to gray matter changes and investigate the extent to which they correlate with APOE ε4. Further, given that majority of the studies involved older adults, it is difficult to conclude on the relationship between APOE ε4 and ventricular changes in other age groups i.e. young and middle age adults. Besides, involving only older adults gives the notion that by the virtue of their age, they tend to have more expansion of the ventricular system which may be suggested to mask the effect of APOE ε4. Further studies with longitudinal focus enrolling large sample size and involving a wider age group from childhood to older adulthood may therefore be necessary to ascertain which of these two factors (increasing age or the presence of APOE ε4) tend to exert a stronger deleterious effect on ventricular expansion. Further, as there is less support for the combination of ventricular volume and CSF Aβ42 for differentiating MCI and AD from HCs at baseline, future systematic review may consider finding out what the outcome might be when gray matter volumes are combined with CSF biomarkers at baseline. We found that most studies did not investigate the interrelationships between ventricular volume alterations and neuropsychological measures. In future studies, the inclusion of correlational analyses between these variables in both cross-sectional and longitudinal studies should be considered important to provide more clarity on how progressive cognitive impairment relate with brain atrophy. The current study did not focus on how APOE ε4, CSF biomarkers, and neuropsychological measures interact with each other. The investigation of this interaction which is devoid of neuroimaging techniques may be another interesting area in future systematic review, as evidence of significant correlations between these variables may be potentially useful biomarkers in the absence of imaging modalities, especially in low-resource settings. Then, a comparison can be made between this outcome and when the variables are combined with validated neuroimaging techniques for AD (MRI and/or PET). Another interesting area to explore may be to shift focus to investigative the interaction of plasma biomarkers with APOE ε4 and neuropsychological measures, and compare with the use of CSF biomarkers.

### 4.3. Conclusion

An important highlight is that there is progressive ventricular volume increase with severity observed in patients with AD, and this is suggested to occur annually at an average volume of 4.4 – 4.7 cm^3^ for AD patients compared to the average volumes of 2.7 – 2.9 cm^3^/year and 1.1 – 1.4 cm^3^/ year for patients with MCI and cognitively normal individuals respectively. The mean change of the ventricular volume for progression from MCI to AD is estimated to be 1.7 cm^3^/year, however further studies are required to confirm this. We further highlight that APOE ε4 is an independent risk factor for the enlargement of the cerebral ventricles in aging and dementia, and the presence and effect of APOE ε4 homozygosity should be considered as an important factor that may modulate increased ventricular expansion over time. Importantly, greater effect of APOE ε4 is exerted on the ventricular system in individuals with dementia, with AD patients being the most affected, and the lateral ventricle being the most affected region. Hence the lateral ventricle may be a potentially useful biomarker in the AD continuum. At baseline and follow-up, we found that the results of the relationship between cerebral ventricular volume and CSF biomarkers showed a stronger preference for Aβ42 compared to tau. Although, the ability of the combination of ventricular volume and Aβ42 for differentiating MCI and AD from HCs seems not to be robust at baseline, the combination at follow-up was more robust, suggesting that the combination may suffice for predicting and tracking the progression of different types of dementia, and more favourably for patients within the clinical stages of MCI and AD. The combination of ventricular volume with MMSE scores showed the most robust performance for differentiating AD and MCI from HCs at both baseline and follow-up. Although the combination of ventricular volume with either ADAS-cog or CDR-sum may also prove as useful biomarkers for tracking the progression of AD over time, there is a stronger preference for the use of MMSE.

## Data Availability

All extracted data are included in the manuscript.

## Funding

None

## Notes

### Competing Interest Statement

The authors have declared no competing interest.

### Funding Statement

No funding was received for the present work.

### Author Declarations

This is a systematic review.

